# Infectious and Clinical Tuberculosis Trajectories: Bayesian modeling with case finding implications

**DOI:** 10.1101/2022.06.27.22276965

**Authors:** Theresa S Ryckman, David W Dowdy, Emily A Kendall

**Author notes:** **Corresponding Author:** Theresa Ryckman.

## Abstract

**Background:** The importance of finding people with undiagnosed tuberculosis (TB) hinges on their clinical and infectious trajectories. Assays for systematic screening should be optimized to find those whose TB will contribute most to future transmission or morbidity.

**Methods:** We constructed a mathematical model which tracks the disease trajectories of individuals with TB, classifying them over time by bacterial burden (smear positive/negative) and symptom status (symptomatic/subclinical). We used Bayesian methods to calibrate this model to historical survival data and notification, mortality, and prevalence survey data from five countries. We combined the resulting individual disease trajectories with evidence on infectiousness, to compare how much different subsets of prevalent TB contribute to future transmission events.

**Results:** Nearly all (89% [95% uncertainty range 83-93%]) smear-negative subclinical TB resolved before diagnosis or treatment, typically after a short disease course (4.3 [3.3-6.7] months). In contrast, people with smear-positive subclinical TB had a longer overall duration of undiagnosed disease (15.5 [11.0-21.3] months), and most eventually developed symptoms. Despite accounting for only 11-20% of prevalent disease, smear-positive subclinical TB accounted for 37-48% of future transmission – a greater contribution than symptomatic TB or smear-negative TB.

**Conclusions:** Subclinical TB with a high bacterial burden accounts for a disproportionate share of future transmission. Priority should be given to developing inexpensive, easy-to-use assays for screening both symptomatic and asymptomatic individuals at scale – akin to rapid antigen tests for other diseases – even if these assays lack the sensitivity to detect paucibacterial disease.

## Introduction

Tuberculosis (TB) remains among the leading causes of global mortality. One likely driver of TB’s persistent global impact is the high burden of undiagnosed TB. Prevalence surveys reveal multiple person-years lived with TB for each person who is officially diagnosed (1). This undiagnosed TB varies in its infectiousness and symptoms (2). In particular, a large fraction of prevalent TB is subclinical: 36-80% of people with TB will not be classified as symptomatic by a typical symptom-screening questionnaire (3). Even among people whose TB has a high enough bacterial burden to be detectable by sputum smear microscopy (a minority of prevalent disease, and an indicator of high transmissibility), 34-68% lack symptoms (4). Thus, a substantial portion of prevalent TB may be highly infectious but would not be detected by the routine health system or by symptom-based screening approaches.

Systematic screening, or active case finding, has been recommended by global bodies in response to the high burden of undiagnosed TB (5, 6). However, the tests currently available for TB screening have important limitations. Current screening tests recommended by World Health Organization guidelines either miss subclinical TB (in the case of symptom screening) or are resource-intensive in ways that limit accessibility (5). Most screening tests also fall short of the target sensitivity of 90% set by existing target product profiles (5, 7). Rethinking priorities for screening tests, and considering less sensitive tests that are optimized on other characteristics, might have logistical or cost advantages.

The importance of finding TB in people with no current symptoms depends on both their current infectiousness and their future course of infectiousness, symptoms, and care-seeking. For people whose TB will soon be treated, resolve without treatment, or remain minimally infectious and asymptomatic, early diagnosis may have little impact. But for people whose TB could make a large cumulative contribution to transmission – due to the combination of their infectiousness and the total time they will spend with undiagnosed TB – the benefits of early detection could be substantial.

Because TB is typically treated once diagnosed, there are few direct data on how disease progresses over time. Epidemiological modeling can address this limitation by synthesizing evidence across data sources to draw longitudinal inferences. We sought to use such a modeling approach to describe the future clinical and infectious trajectories of prevalent TB and thus identify efficient screening approaches. Focusing on HIV-negative adults for the sake of tractability and data alignment, we integrated a wide variety of data sources – including survival data from historical cohorts and recent prevalence surveys from five countries – in a model of the bidirectional evolution of both TB symptoms (subclinical versus symptomatic) and sputum bacterial burden (smear-positive versus smear-negative). By estimating how much different forms of prevalent TB will contribute to transmission and adverse clinical outcomes in the future, the results of this model can inform research and development for TB screening tools.

## Methods

We developed a Markov state-transition model to characterize the expected trajectories of prevalent bacteriologically positive TB disease at a population level. For simplicity and consistency across data sources, we focused on HIV-negative adults with no previous history of TB treatment. We categorized TB disease into four stages, based on a time-dependent classification of whether an individual would endorse TB symptoms on a standard symptom screen (i.e., subclinical versus symptomatic) and whether microscopy of an expectorated sputum specimen would be positive for acid-fast bacilli if performed (i.e., smear-negative versus smear-positive; selected as a proxy for low versus high bacterial burden that is frequently measured in prevalence surveys) (Figure 1). TB was assumed to begin from a smear-negative, subclinical state; that is, individuals transition through a smear-negative state before becoming smear-positive, and through a subclinical state before developing symptoms. Similarly, spontaneous resolution of TB (to a non-infectious, asymptomatic, bacteriologically negative state) was assumed to occur only from the smear-negative, subclinical state. We assumed that only those with symptoms can be offered TB treatment (under the standard of care, in which people must first seek care for symptoms) or experience death due to TB.

**Figure 1:**
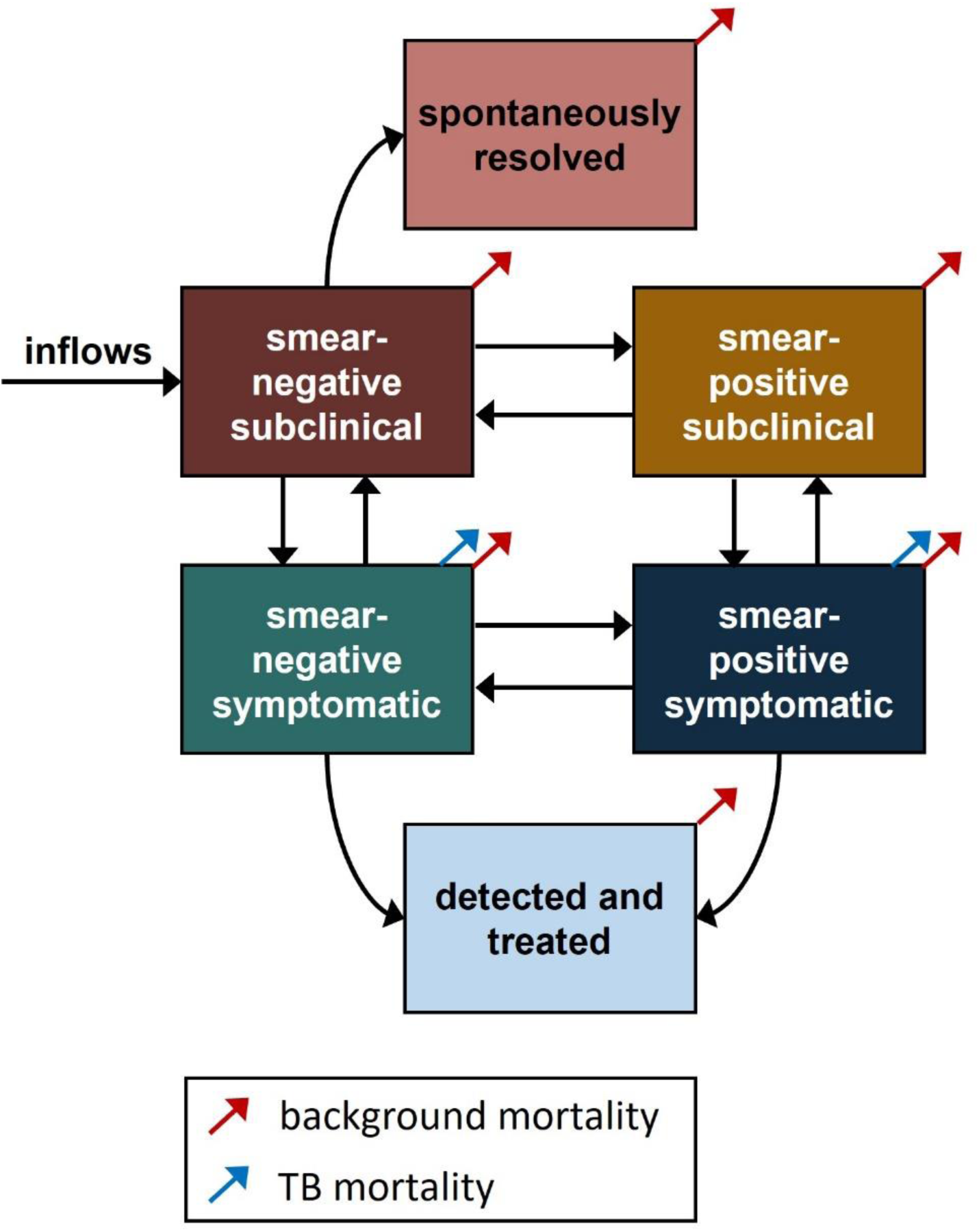
TB natural history model. Shown is the Markov state-transition structure used to evaluate clinical and infectious trajectories of tuberculosis (TB) disease. Individuals with TB were grouped according to symptom status (subclinical versus symptomatic) and sputum smear status (smear-positive versus smear-negative), as described in the main text. Boxes represent different TB-related states, while arrows represent transitions between states and in (incident TB cases; modeled in calibration to present-day targets only, in order to reach a steady state) and out (TB and non-TB deaths) of the model.

The model progresses in monthly time steps, with monthly probabilities of symptom progression, symptom regression, smear status progression, smear status regression, spontaneous resolution, treatment, non-TB (“background”) mortality, and TB-specific mortality. Individuals who resolve spontaneously leave the model (though we evaluated in sensitivity analysis the possibility of recrudescence from the spontaneously resolved state), as do individuals who initiate treatment (as they are no longer treatment-naïve). To induce correlations between smear and symptom status, we assumed in the main analysis that the relative risk of progressing in smear status if already symptomatic equals the relative risk of developing symptoms if already smear-positive, and likewise the relative risk of a smear-positive symptomatic individual regressing in smear status equals the relative risk of them clearing their symptoms; this assumption was also evaluated in sensitivity analysis. All details and model equations are presented in the Appendix, and reproducible model code is available at github.com/rycktessman/tb-natural-history. Analyses were performed in R version 4.0.3 (R Foundation for Statistical Computing, Vienna, Austria).

### Data and Setting

All transition probabilities in the model (other than background mortality) were calibrated to data from the scientific literature (Table 1, full details in the Appendix). These data included historical evidence on TB survival in the pre-antibiotic era and present-day data on TB prevalence, notifications, and mortality in Bangladesh, Cambodia, Nepal, the Philippines, and Vietnam (8–13). These countries were selected because they have: (a) high TB incidence (> 100 per 100,000 per year according to WHO estimates); (b) recent TB prevalence surveys with reporting of smear and symptom status; and (c) low prevalence of HIV coinfection and TB drug resistance (factors likely to alter disease trajectories but not differentiated in our model). We expect inferences about the natural course of disease to generalize to HIV-negative adults in other settings as well. In our model, cross-sectional prevalence data correspond to the distribution of TB across smear and symptom states under current diagnosis and treatment practices. Notification and mortality data correspond to transitions out of the model via treatment or death (with additional unobserved exits from the model via spontaneous resolution or unnotified treatments). Historical survival data consisted of reviews of TB cohort studies from the pre-antibiotic era (14, 15), from which we extracted five- and ten-year survival probabilities after being diagnosed with symptomatic TB, stratified by smear status at the time of diagnosis and pooled across cohorts using Poisson meta-regression.

**Table 1:** Model parameters, prior distributions, and calibration targets.

| **Calibration targets (Philippines)** | **Value** | **Source** |
| --- | --- | --- |
| Percent of prevalent TB that is smear-positive | 37% [32-42%] | Philippines 2016 Prevalence Survey (9) |
| Percent of prevalent TB that is symptomatic | 29% [24-34%] | Philippines 2016 Prevalence Survey (9) |
| Percent of prevalent TB that is smear-positive and symptomatic | 17% [13-23%] | Philippines 2016 Prevalence Survey (9) |
| Ratio of smear-positive prevalence to smear-positive notifications | 1.95 [1.05-3.10] | Estimated – details in appendix |
| Unnotified TB mortality per prevalent case | 2.0% [0.1-3.1%] | Estimated – details in appendix |
| Percent of TB notifications that are smear-positive | 45% [32-62%] | Estimated – details in appendix |
| **Calibration targets (Vietnam)** | **Value** | **Source** |
| Percent of prevalent TB that is smear-positive | 23% [18-29%] | Vietnam 2018-19 Prevalence Survey (13) |
| Percent of prevalent TB that is symptomatic | 36% [29-42%] | Vietnam 2018-19 Prevalence Survey (13) |
| Percent of prevalent TB that is smear-positive and symptomatic | 11% [8-16%] | Vietnam 2018-19 Prevalence Survey (13) |
| Ratio of bacteriologically positive prevalence to notifications | 3.9 [3.0-4.8] | Estimated – details in appendix |
| Unnotified TB mortality per prevalent case | 4.4% [1.9-6.9%] | Estimated – details in appendix |
| Percent of TB notifications that are smear-positive | 68% [54-82%] | Estimated – details in appendix |
| **Calibration targets (Cambodia)** | **Value** | **Source** |
| Percent of prevalent TB that is smear-positive | 33% [28-38%] | Cambodia 2011 Prevalence Survey (11) |
| Percent of prevalent TB that is symptomatic | 30% [25-35%] | Cambodia 2011 Prevalence Survey (11) |
| Percent of prevalent TB that is smear-positive and symptomatic | 14% [11-18%] | Cambodia 2011 Prevalence Survey (11) |
| Ratio of smear-positive prevalence to smear-positive notifications | 1.3 [0.9-1.7] | Estimated – details in appendix |
| Unnotified TB mortality per prevalent case | 3.6% [1.1-6.0%] | Estimated – details in appendix |
| Percent of TB notifications that are smear-positive | 89% [78-95%] | Estimated – details in appendix |
| **Calibration targets (Nepal)** | **Value** | **Source** |
| Percent of prevalent TB that is smear-positive | 30% [24-36%] | Nepal 2018-19 Prevalence Survey (12) |
| Percent of prevalent TB that is symptomatic | 27% [21-33%] | Nepal 2018-19 Prevalence Survey (12) |
| Percent of prevalent TB that is smear-positive and symptomatic | 11% [8-16%] | Nepal 2018-19 Prevalence Survey (12) |
| Ratio of bacteriologically positive prevalence to notifications | 4.7 [3.5-6.1] | Estimated – details in appendix |
| Unnotified TB mortality per prevalent case | 19.2% [8.4-31.9%] | Estimated – details in appendix |
| Percent of TB notifications that are smear-positive | 73% [55-89%] | Estimated – details in appendix |
| **Calibration targets (Bangladesh)** | **Value** | **Source** |
| Percent of prevalent TB that is smear-positive | 39% [33-45%] | Bangladesh 2015-16 Prevalence Survey (8) |
| Percent of prevalent TB that is symptomatic | 38% [33-44%] | Bangladesh 2015-16 Prevalence Survey (8) |
| Percent of prevalent TB that is smear-positive and symptomatic | 19% [15-24%] | Bangladesh 2015-16 Prevalence Survey (8) |
| Ratio of bacteriologically positive prevalence to notifications | 3.9 [3.2-4.5] | Estimated – details in appendix |
| Unnotified TB mortality per prevalent case | 17.1% [8.5-26.2%] | Estimated – details in appendix |
| Percent of TB notifications that are smear-positive | 92% [79-98%] | Estimated – details in appendix |
| **Historical calibration targets (all countries)** | **Value** | **Source** |
| 5-year mortality of smear-negative symptomatic TB | 13% [9-16%] | Pooled estimate of studies in (14, 15) |
| 10-year mortality of smear-negative symptomatic TB | 21% [15-26%] | Pooled estimate of studies in (14, 15) |
| 5-year mortality of smear-positive symptomatic TB | 58% [51-64%] | Pooled estimate of studies in (14, 15) |
| 10-year mortality of smear-positive symptomatic TB | 71% [65-77%] | Pooled estimate of studies in (14, 15) |
| **Prior parameter distributions (all countries)** | **Distribution [lower bound, upper bound]** | **Notes/Rationale** |
| Monthly progression probability from smear-negative subclinical to smear-positive subclinical | Uniform [0%, 20%] | Minimally informative* |
| Monthly progression probability from smear-negative subclinical to smear-negative symptomatic | Uniform [0%, 20%] | Minimally informative* |
| Relative risk of smear (symptom) progression if already symptomatic (smear-positive) | Uniform [1, 10] | Constrained to be $\geq 1$ |
| Monthly regression probability from smear-positive symptomatic to smear-negative symptomatic | Uniform [0%, 20%] | Minimally informative* |
| Monthly regression probability from smear-positive symptomatic to smear-positive subclinical | Uniform [0%, 50%] | Upper bound of 50% corresponds to a symptom duration of $\geq 2$ weeks in the average person with symptomatic TB |
| Relative risk of smear (symptom) regression if subclinical (smear-negative) | Uniform [0, 1] | Constrained to be $\leq 1$ |
| Monthly probability of spontaneous cure from smear-negative subclinical | Uniform [0%, 50%] | Upper bound of 50% was selected to align with that of symptom regression |
| Monthly probability of treatment from smear-negative symptomatic | Uniform [0%, 25%] | Minimally informative* |
| Relative risk of being treated if smear-positive symptomatic (vs. smear-negative symptomatic) | Uniform [1, 20] | Constrained to be $\geq 1$ |
| Monthly probability of TB mortality from smear-negative symptomatic | Uniform [0%, 10%] | Minimally informative* |
| Relative risk of TB mortality if smear-positive symptomatic (vs. smear-negative symptomatic) | Uniform [1, 20] | Constrained to be $\geq 1$ |
| **Non-calibrated transition probabilities** | **Value** | **Source** |
| Annual all-cause mortality (present-day, Philippines) | 0.5% | WHO life tables for adults in each country (25) |
| Annual all-cause mortality (present-day, Vietnam) | 0.4% |  |
| Annual all-cause mortality (present-day, Cambodia) | 0.7% |  |
| Annual all-cause mortality (present-day, Nepal) | 0.5% |  |
| Annual all-cause mortality (present-day, Bangladesh) | 0.4% |  |
| Annual all-cause mortality (historical) | 1% | Based on (15, 26) |
| **Other non-calibrated parameters** | **Value** | **Source** |
| Relative infectiousness of smear-negative TB (vs. smear-positive with the same symptom status) | 35% | Based on (27–29); details in appendix |
| Relative infectiousness of subclinical TB (vs. symptomatic with the same smear status) | 70% | Based on (30–38); details in appendix |

Brackets show 95% confidence intervals around the calibration target means. All targets and estimated values are restricted to treatment-naïve individuals, as detailed in the appendix.
\* The specified upper bound was selected for efficiency, after initial calibration runs were not found to prefer values above the specified range.

### Calibration

We calibrated the model to historical data and each country’s present-day data using Bayesian Incremental Mixture Importance Sampling (16) with minimally informative, uniformly distributed priors (Table 1). Separate historical and present-day simulations, with shared natural history parameters but distinct treatment assumptions and calibration targets, contributed to a single likelihood for each country (details in Appendix).

### Estimation of Future Clinical Trajectories

After calibrating model parameter values to observed data for each country, we simulated individual trajectories for a cohort of 100,000 people with prevalent TB. The cohort’s initial state corresponded to the distribution of symptoms and smear status in each prevalence survey. For each set of parameter values in the calibrated posterior distribution, we ran the model forward in time for five years. We characterized the resulting trajectories in terms of (a) the proportion of individuals in each starting symptom/smear state who ever reached other specified states and (b) the mean cumulative time spent in each state (and with active TB of any kind), averaged across all individuals starting in a given state. Both outcomes are presented as means, with uncertainty intervals corresponding to the 2.5^th^ and 97.5^th^ percentiles, across all posterior parameter sets. Means and uncertainty intervals were calculated for each country separately (to generate country-specific estimates of outcomes) and for all countries combined (to generate pooled estimates of outcomes).

### Estimation of Future Infections

The number of future secondary infections expected to arise from a person with TB depends not merely on the infectiousness of their current state but on both the infectiousness and the duration of all TB states they will inhabit before resolution, cure, or death. For example, individuals who begin in a state that is less infectious, but that is likely to persist undetected for a long time or progress to greater infectiousness, could generate more secondary infections than those who begin in a highly infectious state that will quickly lead to diagnosis and/or mortality. Therefore, we calculated the relative contribution to future transmission from individuals beginning in each of the four prevalent TB states in our model. This calculation combined the time spent in each TB state in the simulated disease trajectories with evidence from the literature (e.g., household contact studies) on the relative infectiousness of each state per unit time. Specifically, we estimated that smear-negative TB is 35% as infectious as smear-positive TB when controlling for symptoms, and that subclinical TB is 70% as infectious as symptomatic TB when controlling for smear status (Table 1; details in Appendix). Combining this per-time relative infectiousness with average disease trajectories for each state allowed us to estimate the relative benefits of identifying and successfully treating one additional person with prevalent TB in each state. Weighting the results by the relative size of the population in each state in corresponding prevalence surveys, we also described the country-specific fractions of future transmission likely to arise from individuals currently in each of the four states.

### Sensitivity Analysis

We tested the sensitivity of results to various structural and parameter assumptions, using data from the Philippines (details in Appendix). Additionally, given wide variation in relative infectiousness estimates reported in the literature and potential biases in interpreting the literature estimates as instantaneous relative infectiousness, we assessed the sensitivity of the simulation results to the relative infectiousness parameters.

## Results

### Calibration

In nearly all calibrated simulations, bacterial status was more stable than symptom status (Figure 2). For example, the estimated monthly probabilities of smear progression (ranging from mean 2.1% [95% CI 1.4-3.0%] in Vietnam to 3.9% [2.8-5.3%] in Bangladesh) and regression (from 1.5% [0.4-3.7%] in Nepal to 1.9% [0.3-6.3%] in Bangladesh) for those with subclinical TB were substantially lower than the corresponding probabilities of symptom progression (from 9% [6-15%] in the Philippines to 17% [12-20%] in Bangladesh) and regression (from 35% [20-49%] in the Philippines to 43% [33-50%] in Bangladesh) among those with smear-negative TB. Among those with smear-positive symptomatic TB, diagnosis and treatment occurred much more frequently (≥7% per month in 95% of parameter sets for all countries) than smear regression (<2% per month in 95% of parameter sets); the rate of symptom regression was less constrained by the calibration process and ranged from <1% to 42% per month. The probability of spontaneous resolution from the smear-negative subclinical state was >20% per month in all countries (in >95% of calibrated simulations). Calibrated models fit closely to both historical and contemporary data (Figures E2-3), and the posterior distributions of most parameters were similar across countries (Figure 2).

**Figure 2:**
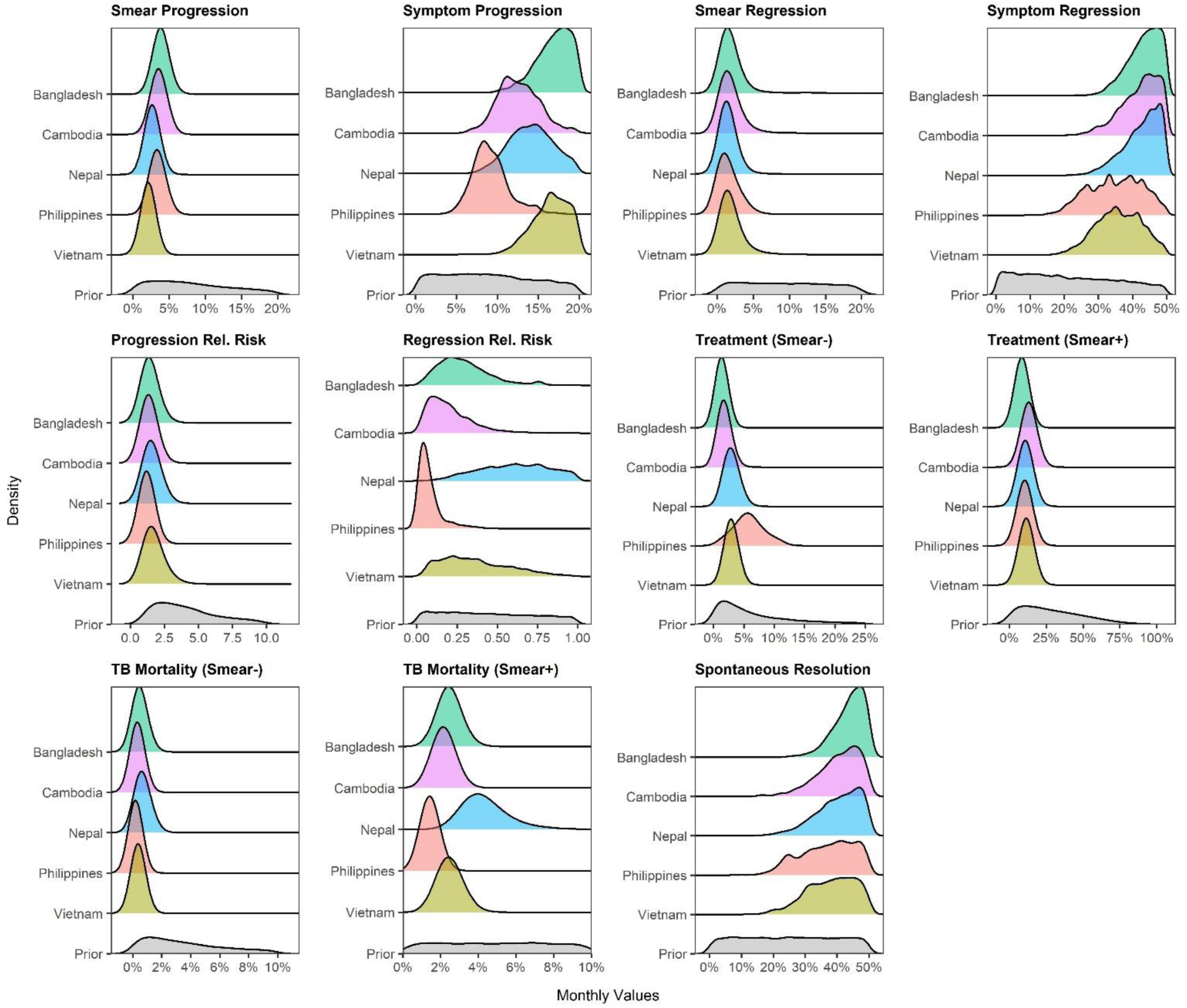
Results of model calibration: posterior parameter distributions. Grey regions show the prior distributions for each parameter, whereas the colored curves show the best-fitting (posterior) distributions after model calibration for each country. Values on the x-axis represent monthly probabilities, except for the two relative risk parameters. Progression relative risk refers to the increased risk of smear or symptom progression when one is symptomatic or smear-positive, compared to being subclinical or smear-negative, respectively. Regression relative risk refers to the decreased risk of smear or symptom regression when one is symptomatic or smear-positive, compared to being subclinical or smear-negative, respectively. Treatment and TB mortality transitions are only shown by smear status because these were assumed only to occur in the presence of symptoms. All priors were sampled from uniform distributions, but exclusion of infeasible parameter sets (for which the transitions out of a state exceeded 100%) resulted in some loss of uniformity (see appendix). The shift from relatively flat prior (grey) distributions to narrower posterior (colored) distributions indicates that only certain parameter values were consistent with the data used to inform the calibration process.

### Individual Clinical Trajectories

Among people with prevalent TB pooled across the five countries, those with smear-positive subclinical TB had the longest projected duration of disease: mean 15.5 months [11.0-21.3], most of which would be spent smear-positive (15.0 months [8.7-22.7]) and over half of which would be spent smear-positive and subclinical (8.5 months [4.6-13.2]) (Figure 3). In addition, 94% [81-99%] of these individuals with smear-positive subclinical TB were projected to eventually develop symptoms; ultimately, 69% [47-85%] would be treated and 16% [8-28%] would die of TB. In contrast, 89% [83-93%] of those with smear-negative subclinical TB were projected to resolve, with an average duration of only 4.3 [3.3-6.7] months with TB, and with only 10% [6-15%] of individuals ever becoming smear-positive. Although the expected TB disease duration of those with smear-positive symptomatic TB (mean 10.9 [6.9-15.6 months]) was also longer than that of the smear-negative states due to less spontaneous resolution, it was shorter than that of smear-positive subclinical TB because of earlier case detection. Projected clinical trajectories were generally similar across countries (Figures E4-5).

**Figure 3:**
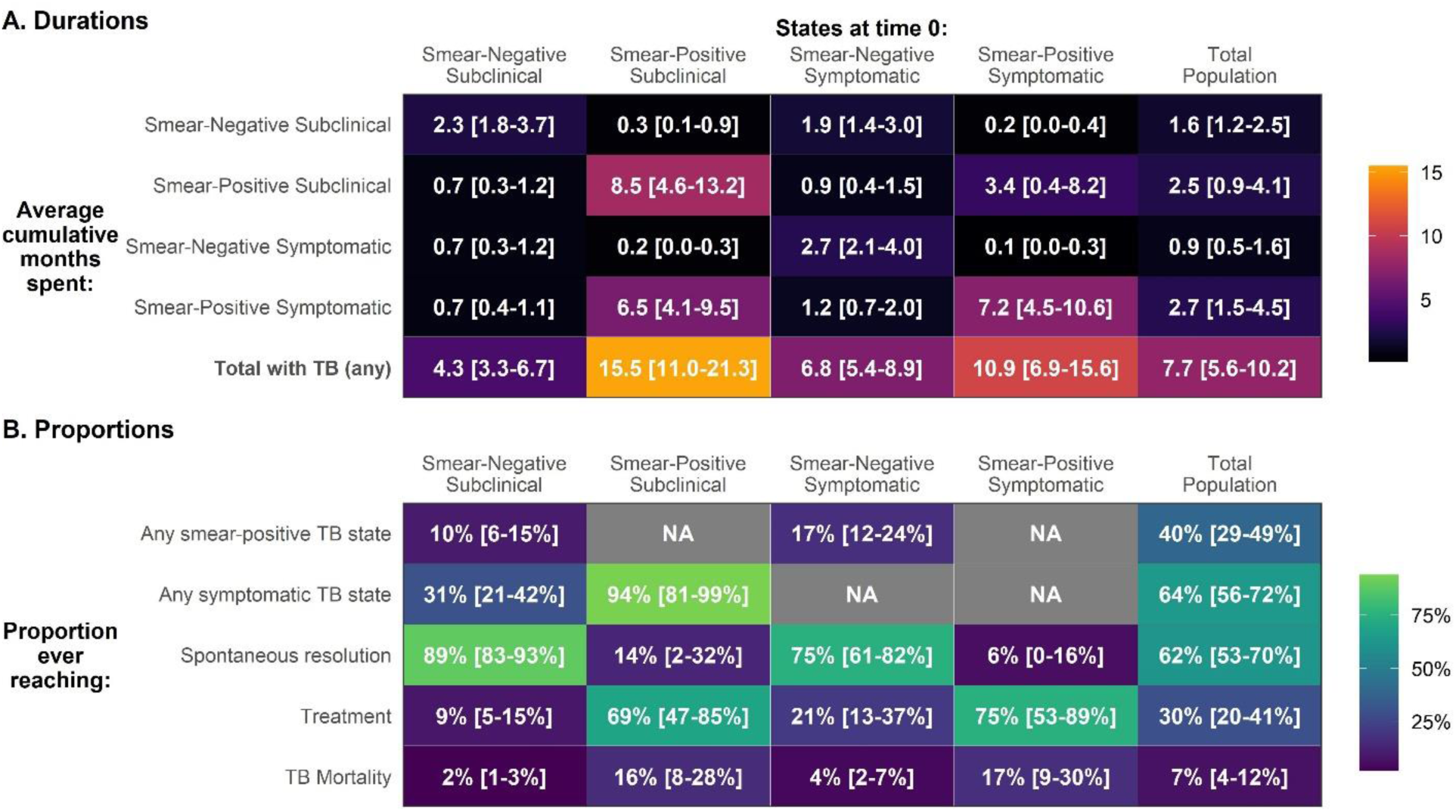
Individual TB natural history trajectory characteristics over five years. Panel A shows the average cumulative durations (in months) spent in each specified state (rows), conditional on being in each starting state (columns) at time 0. Panel B shows the proportion of those in each starting state at time 0 (columns) ever reaching other states/outcomes (rows). Uncertainty intervals for time spent in each state show the 2.5^th^ and 97.5^th^ percentiles across mean values from 50,000 posterior parameter sets for each country pooled together. Uncertainty intervals for the proportions ever reaching a state show the 2.5^th^ and 97.5^th^ percentiles across 50,000 posterior parameter sets for each country pooled together. In both panels, “Total Population” indicates the average times/proportions in a population consistent with each country’s prevalence survey (pooled across the five countries).

### Trajectories of Infectiousness

After incorporating estimates of the relative infectiousness of each state, we estimated that 6.7 [4.4-9.4] times as many secondary infections would arise over the next five years from the average person with current smear-positive subclinical TB, compared to the average person with current smear-negative subclinical TB (Figure 4a). Results varied little between countries (Table E2). Corresponding estimates of other states’ relative contributions to future transmission (continuing to use smear-negative subclinical TB as a reference) were 1.7 [1.4-2.1] times higher for smear-negative symptomatic TB and 5.2 [3.2-7.6] times higher for smear-positive symptomatic TB. If comparing to smear-positive symptomatic TB as a reference, the total number of future infections arising from an individual with current smear-positive subclinical TB was projected to be 1.3 [1.1-1.7] times greater. This finding – that more future infections were estimated to arise from a person with current subclinical smear-positive TB than from a person with current symptomatic smear-positive TB – was robust across all countries and as long as subclinical TB was at least 17% as infectious as symptomatic TB (Figure 4b).

**Figure 4:**
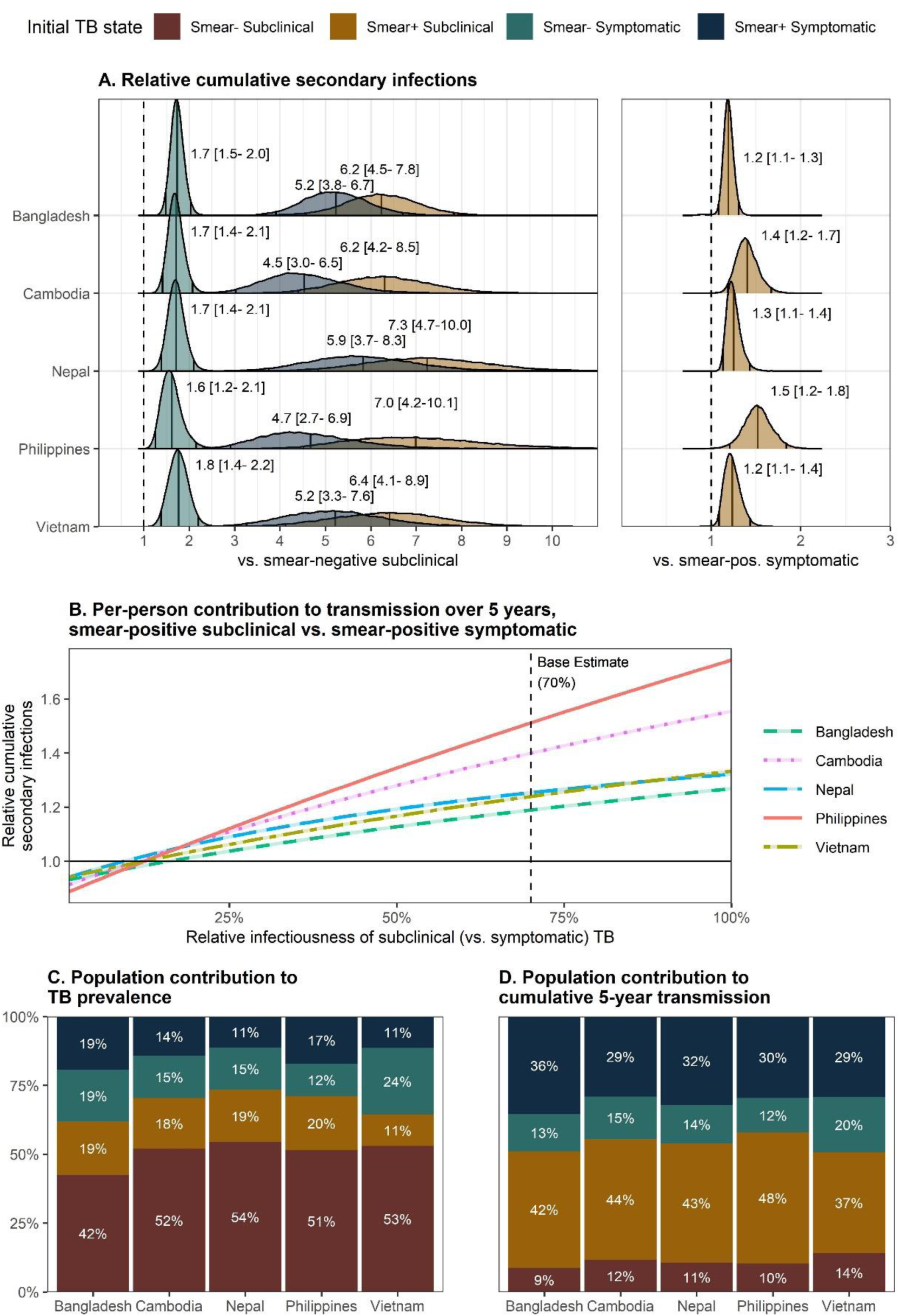
Relative number of future transmission events from individuals with undiagnosed prevalent TB in different symptom and smear states. Both panel A and panel B show the relative cumulative infections generated over 5 years by a single individual in a given TB state at the start of that 5-year period. Panel A shows the full distribution of relative cumulative infections, with vertical lines representing means and 2.5^th^ and 97.5^th^ percentiles across posterior parameter sets. In the left side of panel A, the comparator is someone who initially has smear-negative subclinical TB. In the right side of panel A and in panel B, the comparator is someone who initially has smear-positive symptomatic TB. In panel B, mean relative cumulative infections are compared over a range of values for the per-time relative infectiousness of subclinical (vs. symptomatic) TB. Panels C and D show population-level, rather than individual-level, outcomes: the average contribution of each of the 4 initial TB states to prevalent TB at time 0 (panel C) and to the cumulative transmission generated over the 5 subsequent years by those with prevalent TB at time 0 (panel D).

We calculated population-level contributions to transmission by combining these estimates with the relative prevalence of each state from prevalence surveys. Estimates thus vary by country, and we report the lowest and highest means (and uncertainty intervals) across the five countries. Of all infections generated in the subsequent five years by a cross-sectional sample of people with prevalent TB in the five countries evaluated (i.e., of all transmission potentially avertible by active case-finding at a given moment), we projected that between 37% [31-41%] and 48% [42-53%] would arise from people with current smear-positive subclinical TB, even though they represented only 11-20% of the population with prevalent TB (Figure 4c-d). By comparison, people with current smear-positive symptomatic TB (11-19% of prevalent TB) would generate 29% [23-34%] to 36% [32-39%] of infections, those with current smear-negative symptomatic TB (12-24% of prevalent TB) would generate 12% [10-16%] to 20% [16-25%], and those with current smear-negative subclinical TB (42-54% of the population) would generate the remaining 9% [7-11%] to 14% [10-19%].

### Sensitivity Analyses

In all sensitivity analyses, more secondary infections were estimated to arise from the current smear-positive subclinical state than from any other state (Table E3). Other results differed modestly in some but not all sensitivity analyses. In analyses that excluded 10-year historical mortality targets or allowed for return to prevalent TB after spontaneous resolution, calibrated parameter values (Figure E6) and resulting disease trajectories (Figure E8-9) were similar to the primary analysis. In an analysis that allowed smear and symptom progression/regression to vary independently, progression and regression relative risks were more uncertain, but other parameter distributions remained similar (Figure E6). When limited historical evidence on smear status over time was added to calibration, models matched the main calibration targets less well (Figure E7) but suggested higher TB mortality, more frequent smear and symptom transitions, and somewhat less differentiation in the average durations of different states than in the main analysis (Figures E8-9).

## Discussion

This Bayesian modeling study synthesized historical and contemporary data to determine the future clinical and infectious trajectories of individuals with prevalent undiagnosed TB, according to symptom status and bacterial burden (as classified by symptom screening and sputum smear, respectively). We found that the expected contribution of subclinical disease to future transmission is strongly tied to current bacterial burden – because of corresponding differences not only in current infectiousness, but also in expected disease duration. We projected that individuals with smear-positive subclinical TB – who accounted for only 11-20% of TB in national prevalence surveys – would remain infectious for an additional 13-18 months and generate 37-48% of all future transmission events, depending on the country. By contrast, although smear-negative subclinical disease accounts for a larger proportion of prevalent TB (42-54%), it generated only 9-14% of projected future transmission events, due to a short average TB duration (4.3 months) and high probability of spontaneous resolutions (89%). These results suggest that less than 50% of future transmissions arising from current cases can be averted by active case finding that focuses only on people with symptoms. But if all individuals with high bacterial burdens could be detected regardless of symptoms, then two-thirds or more of transmission could be averted even if those with lower bacterial burdens were missed.

The disproportionate contribution of smear-positive subclinical TB to future transmission has important programmatic implications. Systematic population screening based on symptoms will miss these individuals – but bacteriologic assays need not be highly sensitive to identify them. A target product profile for an assay to identify these individuals – the characteristics of which would resemble rapid antigen tests for COVID-19 – currently does not exist in the TB space. The use case for these assays would be mass screening, not clinical diagnosis. As such, they would differ substantially from current rapid molecular tests, which achieve high sensitivity at the expense of time, complexity, and a cost of $15-30 per test that is unaffordable for mass screening in most low- and middle-income settings (17–19). For an infectivity-focused screening test, sensitivity targets could be relaxed as long as high bacterial burdens were detected – and in turn, more ambitious targets could be set for portability, ease of use, throughput, and cost, with the goal of making widespread systematic screening truly feasible. More research is therefore needed to develop such assays, ideally using more accessible specimens such as exhaled breath, oral fluid, or urine.

A major strength of our model is the integration of a sufficient variety of data sources to simultaneously track symptom and smear status, and to incorporate both progression and regression of each, thus allowing for novel inference about the trajectories of individuals with prevalent undiagnosed TB. Our conclusions were consistent across all sensitivity analyses and five different countries with different TB epidemics and different data limitations, increasing generalizability. Compared to other recent TB modeling analyses, we estimated a similar duration of symptomatic TB but a shorter overall mean duration of TB than Ku et al. (20), likely because modeling spontaneous resolution only from the smear-negative subclinical state allowed such resolution to occur frequently. By contrast, Richards et al. constrained TB disease to last approximately two years (21). Relative to Ragonnet et al. (15), we found a similar rate of smear-negative TB mortality but lower rate of smear-positive mortality; this is likely because we incorporated contemporary as well as historical mortality data, leading to slight underestimates of historical five-year smear-positive mortality (Figure E3). Our finding that smear status is relatively stable suggests that models that treat smear status as fixed (15, 22) are reasonable approximations, at least over shorter-term time horizons.

As with any modeling analysis, this study has certain limitations. First, our Markov model structure did not allow “memory” of an individual’s prior states; in reality, probabilities of disease progression may differ between those who have new incident TB and those who have had more advanced forms previously. Second, we dichotomized symptoms and bacterial burden to correspond with available data, but this dichotomization hides a spectrum. For example, it is possible that the subset of smear-negative TB that is detectable by rapid molecular tests may be more persistent than the average smear-negative TB case, such that detecting Xpert-positive smear-negative TB may yield more benefits than implied here. Dichotomization may also differ between data sources – for example, different prevalence surveys use different definitions of symptomatic, and the average severity of symptoms may be milder (with corresponding lower mortality) among all symptomatic prevalent TB than among the symptomatic patients who sought care and entered historical cohorts. Third, we only included cohort studies of patients with smear-negative TB if they had been smear-positive (or “bacillary”) prior to enrollment, to increase our confidence that the patients being described truly had TB, but it is possible that some of these patients had already experienced spontaneous resolution. Fourth, most evidence on the relationship between smear and symptom status and transmissibility comes from contact investigation and molecular epidemiology studies that are not directly translatable into estimates of instantaneous relative infectiousness and may be subject to biases, for example stemming from behavior change or contact saturation. However, our results were consistent over a wide range of per-time relative infectiousness estimates. Finally, our model is not suitable for representing the population-level dynamics of prevalent TB in high-HIV-burden settings, due to the higher rates of symptom progression and mortality associated with HIV coinfection.

However, our results about individual disease trajectories are expected to apply to HIV-negative adults in settings with both high and low HIV prevalence, as long as those settings have similar passive case detection practices as the countries we modeled.

In summary, a disproportionate amount of future TB transmission likely arises from individuals with undiagnosed smear-positive subclinical TB. To be effective, active case finding strategies must identify this subset of individuals and thus should not rely on symptom screening for triage. By contrast, an estimated 89% of smear-negative subclinical TB may spontaneously resolve, with only minimal contribution to ongoing transmission. Novel tools – for example, lower-sensitivity, high-specificity, low-cost, portable diagnostic assays – to efficiently identify people with smear-positive subclinical TB should therefore be prioritized.

## Data Availability

All data used in the study are available from publicly-available sources cited in the references. No additional data were produced in the present study. Additional model simulation results are available upon reasonable request to the authors.

## Author Contributions

EAK conceptualized the study. TSR curated and cleaned the data, developed the model structure and code, and ran all analyses. TSR wrote the first draft of the manuscript. EAK and DWD contributed to subsequent model drafts and provided oversight and feedback on analyses. EAK and TSR verified the study data. All authors had full access to all the data in the study and the final manuscript and accept responsibility to submit for publication. All authors declare no competing interests.

