## Appendix for "Infectious and Clinical Tuberculosis Trajectories: Bayesian modeling with case finding implications"

**Model Details**

The model assumes that everyone with incident bacteriologically detectable TB starts out smear-negative and subclinical. Each monthly timestep, these smear-negative subclinical individuals can develop a positive smear, develop symptoms, clear their infection without treatment (spontaneously resolve), or die of non-TB causes. Regression and progression of both smear status and symptoms are possible. Returns from the spontaneously resolved state are only considered in sensitivity analysis. Spontaneous resolution is not possible from the smear-positive or symptomatic states; individuals must first regress their smear and/or symptoms. TB mortality and treatment transitions are only possible from the symptomatic TB states, and the probabilities of mortality and treatment are assumed to be higher for smear-positive symptomatic individuals than smear-negative symptomatic individuals.

In analyses that aimed to reach a steady state (calibrating to the present-day prevalence, notification, and mortality targets), deaths re-enter the model as inflows (to the smear-negative subclinical state). In analyses that focused on the future trajectories of those in a particular state (i.e. symptomatic and/or smear-positive), we modeled all individuals as starting in that TB state. This was done when calibrating to the historical cohort data (everyone started out smear-positive symptomatic or smear-negative symptomatic) and when simulating the stability of the different TB states.

**Figure E1: Detailed TB natural history model diagram**

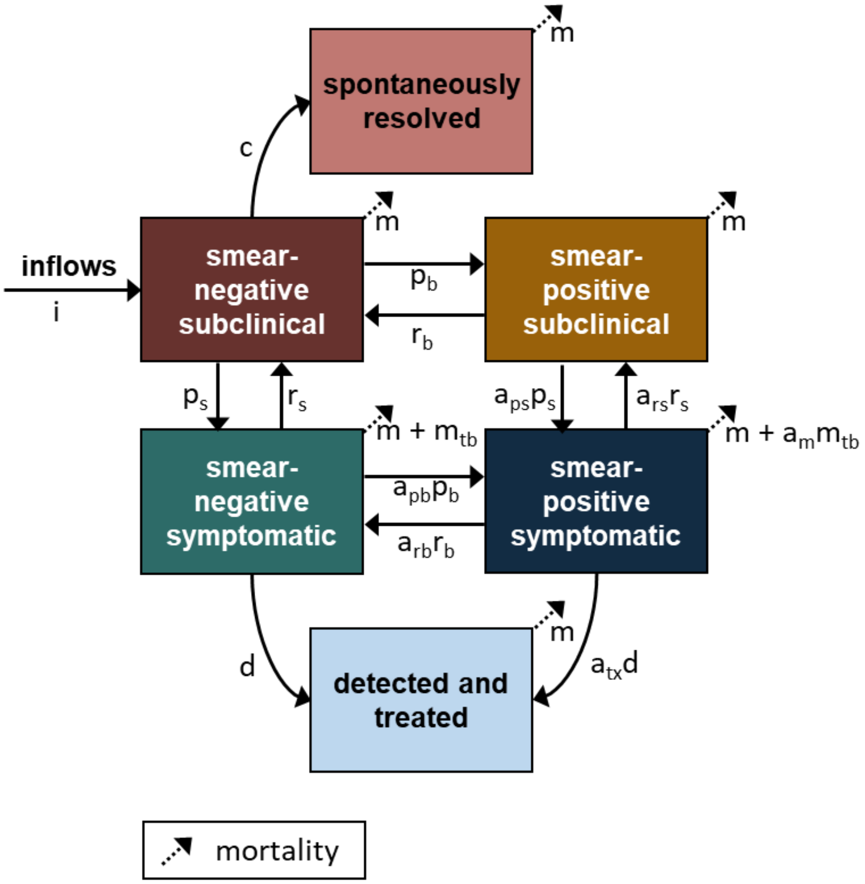

In all analyses, we assumed that the relative risks of smear progression in people with symptoms (vs. without) and of symptom progression in people with smear-positive TB (vs. smear-negative) were greater than one (and similarly, that the relative risks of smear regression and symptom regression for someone who is already symptom- or smear-positive, respectively, are less than one). That is:

$$a_{pb}, a_{ps} > 1$$

$$a_{rb}, a_{rs} < 1$$

In the main analysis, to improve model identifiability and induce and assumed correlation between symptom status and smear status, we further assumed:

$$a_{pb} = a_{ps}$$

$$a_{rb} = a_{rs}$$

That is, the relative risk of progressing in smear status if already symptomatic equals the relative risk of developing symptoms if already smear-positive, and the relative risk of a smear-positive symptomatic individual regressing in smear status equals the relative risk of them clearing their symptoms. These parameters were allowed to vary independently in sensitivity analysis.

*Model Equations*

In the below equations, $s$ refers to symptom status ($s-$ indicates subclinical TB and $s+$ indicates symptomatic TB) while $b$ refers to smear status ($b-$ indicates smear-negative TB and $b+$ indicates smear-positive TB; $b$ was selected to correspond to bacillary load). $Tx$ indicates treated individuals and $Res$ indicates spontaneously resolved individuals. Although the model was implemented with transition probabilities, the differential equation forms of the transitions (using rates) are shown below to allow them to be concise and easier to understand.

$$\frac{dTB_{s-b-}}{dt} = r_b TB_{s-b+} + r_s TB_{s+b-} + i - (c + p_b + p_s + m)$$

$$\frac{dTB_{s-b+}}{dt} = p_b TB_{s-b-} + a_{rs} r_s TB_{s+b+} - \left(r_b + a_{ps} p_s + m\right)$$

$$\frac{dTB_{s+b-}}{dt} = p_s TB_{s-b-} + a_{rb} r_b TB_{s+b+} - (r_s + a_{pb} p_b + d + m_{tb} + m)$$

$$\frac{dTB_{s+b+}}{dt} = a_{ps} p_s TB_{s-b+} + a_{pb} p_b TB_{s+b-} - (a_{rs} r_S + a_{rb} r_b + a_{tx} d + a_m m_{tb} + m)$$

$$\frac{dTx}{dt} = d * TB_{s+b-} + a_{tx} d * TB_{s+b+} - m$$

$$\frac{dRes}{dt} = c TB_{s-b-} - m$$

$$i = m(TB_{s-b-} + TB_{s-b+} + TB_{s+b-} + TB_{s+b+} + Tx + Res) + m_{tb} TB_{s+b-} + a_m m_{tb} TB_{s+b+}$$

### Sensitivity Analyses

To test the sensitivity of results to certain parameter assumptions, we recalibrated the model assuming a 15% (vs. 0%) annual probability of return from the spontaneously resolved state to prevalent infectious TB, comparable to TB incidence in the year after new infection with tuberculin skin test conversion (1) and consistent with estimates from another modeling exercise (2). We also recalibrated a version of the model that decoupled the relative risks of symptom and smear progression and regression, allowing for a total of four independent progression and four independent regression probabilities at the expense of additional model complexity. In addition, we examined sensitivity to the selection of calibration targets. Because our focus is on a five-year timeframe, we explored models that excluded the ten-year historical mortality targets. Another sensitivity analysis incorporated smear status after four years as a calibration target, based on a small subset of historical cohort studies that included information on smear status over time (Table E1). These sensitivity analyses were all conducted using data from the Philippines.

### Calibration Details

*Overview*

We calibrated the Markov model using Incremental Mixture Importance Sampling (IMIS), a Bayesian calibration method that is related to Sampling Importance Resampling but is more computationally efficient (3). For each country we sampled a total of 10.5 million values for each parameter and generated a likelihood-weighted posterior from the 50,000 best-fitting sets of sampled values. To reflect the mix of historical and present-day scenarios that served as sources of calibration data, we performed three distinct analyses under each parameter set for each country to evaluate a joint likelihood. First, to reflect the present-day epidemic, the full model was run with population replacement as an inflow to the smear-negative subclinical state until a steady state was reached (i.e., equilibrium open cohort). Concomitantly, to reflect historical cohort studies, treatment and inflows were turned off, and the model was initialized with either 100% symptomatic smear-positive or 100% symptomatic smear-negative patients (i.e., closed smear-positive and smear-negative cohorts). These latter two cohorts were followed for ten years, and their mortality over time was compared to the historical data on smear-positive and smear-negative mortality, respectively. The joint likelihood was then taken as the product of the likelihoods for each of the three cohorts above, evaluated against their corresponding calibration targets.

*Present-Day Calibration Targets*

Targets used in model calibration are shown in Table 1 in the main text. The proportion of prevalent TB cases that are smear-positive and/or symptomatic were obtained directly from prevalence surveys for the five countries (4–8). In the Philippines prevalence survey, we removed individuals who were currently or had historically been on TB treatment, as this information was directly available in the survey report and our model focuses on a treatment-naïve population. This adjustment was not possible for the other four prevalence surveys, but the proportions who had recently been on or were currently on TB treatment was low. For these countries, we assumed that the distribution of smear status and symptoms was equivalent among treated and among treatment-naïve prevalent TB.

The ratio of smear-positive prevalence to smear-positive notification was obtained directly from the Philippines prevalence survey and was calculated from the Cambodia prevalence survey and the number of smear-positive notifications in Cambodia in 2011 reported to the WHO. The smear-positive prevalence to notification ratio was used rather than the bacteriologically positive prevalence to notification ratio because most bacteriologically-confirmed notifications in the Philippines and Cambodia were diagnosed via smear microscopy at the time of the survey (the Cambodia survey was conducted in 2011, before widespread Xpert rollout, and only 13% of Philippines notifications were tested using Xpert in 2016, regardless of result (9)) and because Cambodia's survey was the only survey conducted during a year in which the smear breakdown of notifications was still reported to the WHO.

In the other three countries, Xpert diagnosis was more common and/or the prevalence to notification ratio of smear-positive TB was not reported; thus the bacteriologically positive prevalence to notification ratio was used instead. This prevalence to notification ratio was calculated as the prevalence reported in the 2017-18 survey (treatment-naïve; i.e., removing cases that were currently on treatment and, when reported, cases that had been treated in the past two years) divided by the number of new pulmonary notifications reported to the WHO in the year of the prevalence survey, adjusting to remove notifications from children (typically <5% of notifications) and an estimate of the clinically-diagnosed notifications that were not truly TB. For the latter, we assumed country-specific percentages of clinically-diagnosed notifications were actually TB. These percentages varied from 34% [23-47%] in Vietnam, where country-specific evidence was available (10) and 25% [4-54%] in Nepal and Bangladesh, where less applicable (or no) country-specific evidence was available and thus a wider range based on evidence from multiple countries was incorporated, including published literature and the proportions of individuals with a positive symptom screen and abnormal chest Xray that had TB in the respective prevalence surveys (7, 8, 10–14). Uncertainty in these inputs to the calibration target was incorporated by sampling from distributions corresponding to each input, calculating the target value implied by these inputs, and computing the mean, 2.5$^{th}$, and 97.5$^{th}$ percentiles of the resulting distribution of target estimates.

The ratio of mortality to prevalence was another calibration target (equations 1-3). For this target, we adjusted estimates of TB mortality from the WHO to remove mortality that occurred while on treatment, since our model focuses on a treatment-naïve population and does not include TB mortality from the detected and treated state. This adjustment was made by subtracting out the proportion of TB notifications whose outcomes were tracked that died, as reported to the WHO. Lower and upper bounds on the treated TB case fatality ratio were based on assumptions made by the WHO, resulting in treated TB case fatality ratio estimates of 2.4% [1-7%] in the Philippines and Vietnam, 2.3% [1-7%] in Cambodia, 2.9% [1-7%] in Nepal, and 3.8% [1-7%] in Bangladesh, as follows: Deaths among notifications whose treatment failed or who were lost to follow-up (based on the WHO-estimated ratio of mortality to incidence) and among treated but unnotified individuals (based on (15) for Vietnam and prevalence survey data on where current TB cases were treated for the other countries) were also estimated and removed from the estimate of untreated deaths. The estimate of untreated deaths was divided by the estimate of treatment-naïve prevalence from the prevalence survey. The ratio of unadjusted deaths to treatment-naïve prevalence was 3.6% [3.0-4.3%] in the Philippines, 5.6% [3.6-8.0%] in Vietnam, 5.2% [3.3-7.3%] in Cambodia, 21.0% [9.8-34.1%] in Nepal, and 20.9% [12.6-30.0%] in Bangladesh. The ratio of adjusted deaths to treatment-naïve prevalence was 2.0% [0.1-3.1%] in the Philippines, 4.4% [1.9-6.9%] in Vietnam, 3.6% [1.1-6.0%] in Cambodia, 19.2% [8.4-31.9%] in Nepal, and 17.1% [8.5-26.2%] in Bangladesh.

$$[1]\ deaths_{untx} = deaths_{WHOest} - \frac{1}{p(notif|tx)}\left(cfr_{treat}notif_{nofailLTFU} + \left(\frac{deaths_{WHOest}}{cases_{WHOest}}\right)notif_{failLTFU}\right)$$

$$[2]\ prev_{untx} = prev * (1 - prop_{txhist})$$

$$[3]\ untx\ mort\ to\ prev\ ratio = \frac{deaths_{untx}}{prev_{untx}}$$

The final present-day calibration target was the proportion of notified [i.e., treated] patients who are smear-positive at the time of notification (equations 4-7). We assumed that all lab-confirmed notifications not diagnosed via Xpert were smear-positive and some Xpert diagnoses and clinical diagnoses were smear-positive; we estimated these from WHO-reported data using equation 4. Countries report to WHO the number of notifications that received a rapid diagnostic (i.e., Xpert), but not the number that tested positive (some Xpert negatives may still be clinically diagnosed). We estimated that 90% of those who received an Xpert test and were subsequently diagnosed with TB would have had a positive Xpert result and thus would be counted as clinical diagnoses, but varied this widely [57-100%], based on studies on the use of Xpert in real-world settings (16, 17). We then estimated that 44% [26-68%] of these Xpert-positive cases would have also been smear-positive (based on studies that reported on smear microscopy results among Xpert positive patients (16, 18, 19)), despite not being tested using smear microscopy, and thus should not be removed from the estimate of notified smear-positive cases. We removed the estimated Xpert-positive smear-negative notifications from the reported lab-confirmed notifications to estimate the number of lab-confirmed notifications that were smear-positive (second line of equation 4). We assumed 0 Xpert diagnoses were made in Cambodia (2011) and Bangladesh (2015).

We also added the estimated number of clinically-diagnosed notifications that would have tested positive on smear microscopy if they had been tested (first line of equation 4). We assumed that the proportion of clinical diagnoses that would test positive on smear microscopy equals the product of the proportion of clinical diagnoses that truly have TB, the proportion of clinical diagnoses that were not tested using smear microscopy (since many clinical diagnoses could include those who tested negative on smear), and the proportion of clinical diagnoses that truly have TB and did not receive a smear test who would be smear-positive if tested (equation 5). The first of these quantities (proportion of clinical diagnoses that truly have TB) was estimated to be anywhere from 10-61% (mean estimate 30%) in the Philippines, where a strong country-specific estimate was lacking (the percent of symptomatic individuals with an abnormal chest Xray from the Philippines prevalence survey was used to parameterize the lower bound of 10%) (4, 10–12). In Cambodia, we estimated that 25% [12-37%] of clinical diagnoses were truly TB, based on country-specific studies and using the percent of individuals with a positive symptom screen and abnormal chest Xray that had TB in the prevalence survey to parameterize the lower bound (14). For the other countries, we used the same estimated percent that was assumed for the calculation of bacteriologically-positive prevalence to bacteriologically-positive notifications (25% [4-54%] in Nepal and Bangladesh and 34% [23-47%] in Vietnam, details above). The second quantity (proportion of clinically diagnoses that did not have a sputum smear microscopy test at all) was estimated from the Philippines prevalence survey (58% [42-74%] in the private sector and 19% [0-38%] in the public sector), which included questions for individuals currently/recently on treatment about where they received care and what diagnostic tests were conducted, and data from Vietnam on smear testing of presumptive TB patients (18% [9-29%]) (10). In Cambodia, Bangladesh, and Nepal we assumed that smear testing was generally widely available (only 10% [5-17%] would not have been smear tested) based on the survey reports and WHO reports on smear availability (20). The third quantity (proportion of clinical diagnoses with non-smear-tested TB that would have been smear-positive if tested) was estimated to range anywhere from the proportion of all TB cases in the prevalence survey that were smear-positive (lower bound) to the upper bound on the proportion of lab-confirmed TB notifications that are smear-positive described in the previous paragraph. This quantity thus also varied slightly by country: 54% [37-71%] in the Philippines, 44% [23-67%] in Vietnam, 55% [33-78%] in Cambodia, 51% [30-73%] in Nepal, and 68% [38-92%] in Bangladesh.

The estimated number of smear-positive notifications was divided by the estimated number of total notifications (equation 7), which was adjusted from reported notifications to remove false-positives (equation 6). This adjustment was made using the same estimated proportion of clinical diagnoses that truly have TB from the previous paragraph (30% [10-61%] in the Philippines, 25% [4-54%] in Nepal and Bangladesh, 34% [23-47%] in Vietnam, and 25% [12-37%] in Cambodia).

[4] $$notif_{smear+} = notif_{lab\ conf} + notif_{clin\ dx} p(smear + |clin\ dx) - notif_{rapid\ test} p\big(Xpert + \big| Xpert\ tested\ \&\ treated\big)\big(1 - p(smear + |Xpert +)\big)$$

[5] $$p(smear + |clin\ dx) = p(TB|clin\ dx)p(no\ smear|clin\ dx)p(smear + |no\ smear, TB, clin\ dx)$$

[6] $$notif_{TB} = notif_{lab\ conf} + notif_{clin\ dx} * p(TB|clin\ dx)$$

[7] $$prop_{smear+} = \frac{notif_{smear+}}{notif_{TB}}$$

*Historical Calibration Targets*

Data on the mortality and survival of historical cohorts of TB patients were pooled from studies reported in Tiemersma et al. and Ragonnet et al. (21, 22). We excluded studies from the post-antibiotic era, studies that did not provide estimates of mortality and survival at either five years or ten years after study start, and studies that included patients with suspected TB that was never bacteriologically confirmed (i.e., patients were never observed as having "open TB" or "bacillary TB"). The latter exclusion was made to avoid inclusion of false-positives. Thus all patients in smear-negative cohorts would have been smear-positive at one point. For consistency with prior studies, we classified "open" TB or "bacillary" TB as smear-positive and "closed" TB or "abacillary" TB as smear-negative. We used Poisson meta-regression with random effects to generate a pooled estimate of five-year and ten-year mortality, by smear status (using *xtpoisson* in Stata). Separate regressions were fitted for the smear-positive cohorts and the smear-negative cohorts.

**Table E1: Historical calibration targets used in smear status sensitivity analysis**

| | Value | Source |
|---|---|---|
| Percent of living smear-positive patients still smear-positive at 4 years | 17% [13-21%] | Sinding-Larsen 1937 (23) |
| | 51% [43-58%] | Braeuning & Neisen, reported in Griep 1939 (24) |
| | 25% [14-36%] | Griep 1939 (24) |

*Likelihood Function*

The likelihood of observing the calibration targets given model parameters, $\theta$, and their resulting output, $X(\theta)$ was calculated as the product of the likelihood of observing the present-day targets (using a model with inflows, treatment, and present-day all-cause mortality brought to a steady state; equation 8), the likelihood of observing the historical smear-positive cohort targets (using a model where everyone starts out with smear-positive symptomatic TB and is followed for ten years, with no inflows, no treatment, and historical all-cause mortality; equation 9), and the likelihood of observing the historical smear-negative cohort targets (using a model where everyone starts out with smear-negative symptomatic TB and is followed for ten years, with no inflows, no treatment, and historical all-cause mortality; equation 10).

[8] $like_{present} = like_{smear} * like_{symptom} * like_{smearsymptom} * like_{prevnotifratio} * like_{mortprevratio} * like_{smearnotifprop}$

We assumed likelihoods corresponding to the prevalence survey targets followed binomial distributions. In equations 8a-8c, $\hat{p}$ represents proportions of cases (that are smear-positive, symptomatic, or smear-positive and symptomatic) output by the model and its parameters, $n_{prev}$ is the number of TB cases in the prevalence survey, and $n_{smr}, n_{symp}, n_{smr\,\&\,symp}$ are the numbers of cases in the prevalence survey that were smear-positive, symptomatic, or smear-positive and symptomatic.

[8a] $\mathcal{L}_{smear}(n_{smr}; \hat{p}_{smr}(\theta), n_{prev}) = \binom{n_{prev}}{n_{smr}} (\hat{p}_{smr}(\theta))^{n_{smr}} (1 - \hat{p}_{smr}(\theta))^{n_{prev}-n_{smr}}$

[8b] $\mathcal{L}_{symptom}(n_{symp}; \hat{p}_{symp}(\theta), n_{prev}) = \binom{n_{prev}}{n_{symp}} \left(\hat{p}_{symp}(\theta)\right)^{n_{symp}} \left(1 - \hat{p}_{symp}(\theta)\right)^{n_{prev}-n_{symp}}$

[8c] $\mathcal{L}_{smearsymptom}(n_{smr\,\&\,symp}; \hat{p}_{smr\,\&\,symp}(\theta), n_{prev}) = \binom{n_{prev}}{n_{smr\,\&\,symp}} \left(\hat{p}_{smr\,\&\,symp}(\theta)\right)^{n_{smr\,\&\,symp}} \left(1 - \hat{p}_{smr\,\&\,symp}(\theta)\right)^{n_{prev}-n_{smr\,\&\,symp}}$

The likelihood function for the prevalence-to-notification ratio target was defined using a gamma distribution. Here, the gamma shape, $k$ and scale, $s$, parameters were selected to fit the mean and 95% confidence intervals shown in Table 1 in the main text, and $\widehat{pnr}$ represents the prevalence-to-notification ratio output by the model and its parameters. For the Philippines and Cambodia, the smear-positive prevalence to smear-positive notification ratio was used, and for Vietnam, Nepal, and Bangladesh, the bacteriologically-positive prevalence to bacteriologically-positive notification ratio was used.

[8d] $\mathcal{L}_{pnr}(\widehat{pnr}(\theta); k, s) = \frac{1}{\Gamma(k)s^k} x^{k-1} e^{-\frac{x}{s}}$

For the remaining two present-day targets (untreated deaths to prevalence ratio and the proportion of notifications that are smear-positive), we used empirical distributions that propagated uncertainty in all of the inputs used to estimate these targets (see equations 1-7). This was accomplished by sampling from uncertainty distributions corresponding to each input and calculating the targets for each sampled set of inputs.

For the historical mortality targets, likelihood functions were defined assuming binomial distributions, with parameters $k$ and $n$ fit to the means and 95% confidence intervals from the Poisson meta-regression.

[9] $\mathcal{L}_{hist,smearpos}\left(k_{pos,5}\ k_{pos,10}\,; \hat{p}_{die\ 5,pos}(\theta), \hat{p}_{die10,pos}(\theta), n_{pos,5}, n_{pos,10}\right) = \left[\binom{n_{pos,5}}{k_{pos,5}} \left(\hat{p}_{die5,pos}(\theta)\right)^{k_{pos,5}} \left(1 - \hat{p}_{die5,pos}(\theta)\right)^{n_{pos,5}-k_{pos,5}}\right] * \left[\binom{n_{pos,10}}{k_{pos,10}} \left(\hat{p}_{die10,pos}(\theta)\right)^{k_{pos,10}} \left(1 - \hat{p}_{die10,pos}(\theta)\right)^{n_{pos,10}-k_{pos,10}}\right]$

[10] $\mathcal{L}_{hist,smearneg}\left(k_{neg,5}\ k_{neg,10}\,; \hat{p}_{die\ 5,neg}(\theta), \hat{p}_{die10,neg}(\theta), n_{neg,5}, n_{neg,10}\right) = \left[\binom{n_{neg,5}}{k_{neg,5}} \left(\hat{p}_{die5,neg}(\theta)\right)^{k_{neg,5}} \left(1 - \hat{p}_{die5,neg}(\theta)\right)^{n_{neg,5}-k_{neg,5}}\right] * \left[\binom{n_{neg,10}}{k_{neg,10}} \left(\hat{p}_{die10,neg}(\theta)\right)^{k_{neg,10}} \left(1 - \hat{p}_{die10,negs}(\theta)\right)^{n_{neg,10}-k_{neg,10}}\right]$

[11] $\mathcal{L} = \mathcal{L}_{present} * \mathcal{L}_{hist,smearpos} * \mathcal{L}_{hist,smearneg}$

*IMIS Procedures*

For each country, we randomly sampled 100,000 parameter sets from prior distributions and calculated the likelihood of observing the calibration targets given these parameter sets and their resulting model output. Prior distributions were uniform and relatively uninformed to maximize the influence of calibration targets in informing model parameters. In initial tests of our calibration procedure, posterior parameter distributions did not tend to overlap with the bounds we established on the uniform distributions. The exceptions were symptom regression and spontaneous resolution. We set 50% as the upper bound on the monthly probability of symptom regression, for consistency with prevalence survey definitions of symptoms: most prevalent symptomatic TB cases found in these surveys identify as having had a cough for ≥ 2 weeks. At probabilities at or below 50%, almost all modeled individuals would have symptoms for ≥ 2 weeks. We set an analogous upper bound of 50% on the monthly probability of resolution of smear-negative subclinical TB, thus excluding models in which a large proportion of TB disease episodes have a duration of only 1 month.

IMIS parameterizes a multivariate normal distribution based on the best-fitting parameter sets from the initial random sampling - we sampled 10,000 times from those distributions to identify additional good-fitting parameter sets. This multivariate normal sampling procedure was repeated 10 times. The entire calibration procedure was replicated 50 times, to produce 50 distinct IMIS chains for a total of 5 million random parameter samples and 5.5 million targeted parameter samples for each of the 5 countries. 1000 parameters were sampled with replacement and using sampling weights based on prior and posterior likelihoods for each of the 50 chains, yielding a posterior of 50,000 parameter sets per country. This strategy yielded between 16,783 and 29,380 unique posterior parameter sets in each country. To implement IMIS, we adapted the R package *IMIS* (25).

**Relative Infectiousness Estimates**

Literature-based estimates of the instantaneous relative infectiousness of subclinical TB (compared to symptomatic TB) and smear-negative TB (compared to smear-positive TB) were combined with simulated individual trajectories to infer the relative number of secondary infections generated by individuals and populations in each of the four TB states. Evidence on the relative infectiousness of smear-negative vs. smear-positive TB can be estimated from molecular epidemiology studies or from studies that assessed TB disease incidence or TST/IGRA conversion among household contacts of TB patients.  The instantaneous relative infectiousness of smear-negative TB (compared to smear-positive TB) was estimated as 0.35, based on 5 studies of TB incidence among contacts and 1 study of TST/IGRA conversion among contacts with relative risks ranging from 0.29 to 0.42 (26–31). The estimate of 0.35 is also consistent with evidence from 3 molecular epidemiology studies from the United States, Canada, and the Netherlands (32–34), which estimated relative risks ranging from 0.21 to 0.24, but which may underestimate relative infectiousness in high-burden settings due to (a) assumptions about the attribution of transmission in mixed clusters (clusters where there are multiple possible index cases, some of whom were smear-positive and some of whom were smear-negative) and (b) the lower detection probability of smear-negative cases in settings where smear microscopy is still used to diagnose TB.

Less evidence was available on the relative infectiousness of subclinical vs. symptomatic TB, since most household contact cohort studies and molecular epidemiology studies include mostly (or all) cases with symptoms. The instantaneous relative infectiousness of subclinical TB (compared to symptomatic TB) was estimated as 0.7, based on a study from Cubillos-Angulo et al., which measured TST conversion among household contacts of TB patients, with index patients stratified by whether or not they had cough for greater than 4 weeks at the time of diagnosis (35). This estimate is also reasonably consistent with estimates from 2 household contact TST/IGRA conversion studies that stratified index cases by continuous degree of symptoms (increases in visual analog cough scale or coughs per hour) (36, 37).

We applied the relative infectiousness estimate of subclinical vs. symptomatic TB to both smear-positive and smear-negative cases, and the relative infectiousness estimate of smear-positive vs. smear-negative TB to both subclinical and symptomatic cases. This resulted in estimates that smear-positive subclinical TB is 70% as infectious and smear-positive symptomatic TB, smear-negative symptomatic TB is 35% as infectious as smear-positive symptomatic TB, and smear-negative subclinical TB is 24.5% as infectious as smear-positive symptomatic TB. Because of limited applicability of these estimates to our analysis (see main text, discussion section), we varied these estimates widely in sensitivity analysis.

Sputum Samples among Tuberculosis Suspected Cases in Nyamira County Referral Hospital. *Mycobact Dis* 2017;07:

20. World Health Organization. Global Tuberculosis Report 2020. 2020;at <https://www.who.int/publications-detail-redirect/9789240013131>.
21. Tiemersma EW, van der Werf MJ, Borgdorff MW, Williams BG, Nagelkerke NJD. Natural History of Tuberculosis: Duration and Fatality of Untreated Pulmonary Tuberculosis in HIV Negative Patients: A Systematic Review. *PLoS ONE* 2011;6:e17601.
22. Ragonnet R, Flegg JA, Brilleman SL, Tiemersma EW, Melsew YA, McBryde ES, Trauer JM. Revisiting the Natural History of Pulmonary Tuberculosis: A Bayesian Estimation of Natural Recovery and Mortality Rates. *Clin Infect Dis* 2021;73:e88–e96.
23. Sinding-Larsen M. On the Collapse Treatment of Pulmonary Tuberculosis. A Clinical Adaptation and Follow-up Examination of the Material from Vejlef jord Sanatorium 1906-1932. *Acta Med Scand* 1937;91:7–212.
24. Griep WA. De prognose van de open longtuberculose. 1939;at <https://www.delpher.nl/nl/boeken/view?coll=boeken&identifier=MMUBA08:000000388>.
25. Raftery AE, Bao L. IMIS: Incremental Mixture Importance Sampling. at <https://rdrr.io/cran/IMIS/man/IMIS.html>.
26. Vidal R, Miravitlles M, Caylà JA, Torrella M, Martín N, de Gracia J. [A contagiousness study in 3071 familial contacts of tuberculosis patients]. *Med Clin (Barc)* 1997;108:361–365.
27. Ling D-L, Liaw Y-P, Lee C-Y, Lo H-Y, Yang H-L, Chan P-C. Contact investigation for tuberculosis in Taiwan contacts aged under 20 years in 2005. *Int J Tuberc Lung Dis* 2011;15:50–55.
28. Chan P-C, Peng SS-F, Chiou M-Y, Ling D-L, Chang L-Y, Wang K-F, Fang C-T, Huang L-M. Risk for Tuberculosis in Child Contacts: Development and Validation of a Predictive Score. *Am J Respir Crit Care Med* 2013;131204094545001.doi:10.1164/rccm.201305-0863OC.
29. Gorís-Pereiras A, Fernández-Villar A, Chouciño-Garrido N, Otero-Baamonde M, Vázquez-Gallardo R. Factores predictores de la aparición de nuevos casos de infección tuberculosa y de viraje tuberculínico en un estudio de contactos. *Enferm Clínica* 2008;18:183–189.

**Supplemental Results**

**Figure E2: Fit of main model to calibration targets**

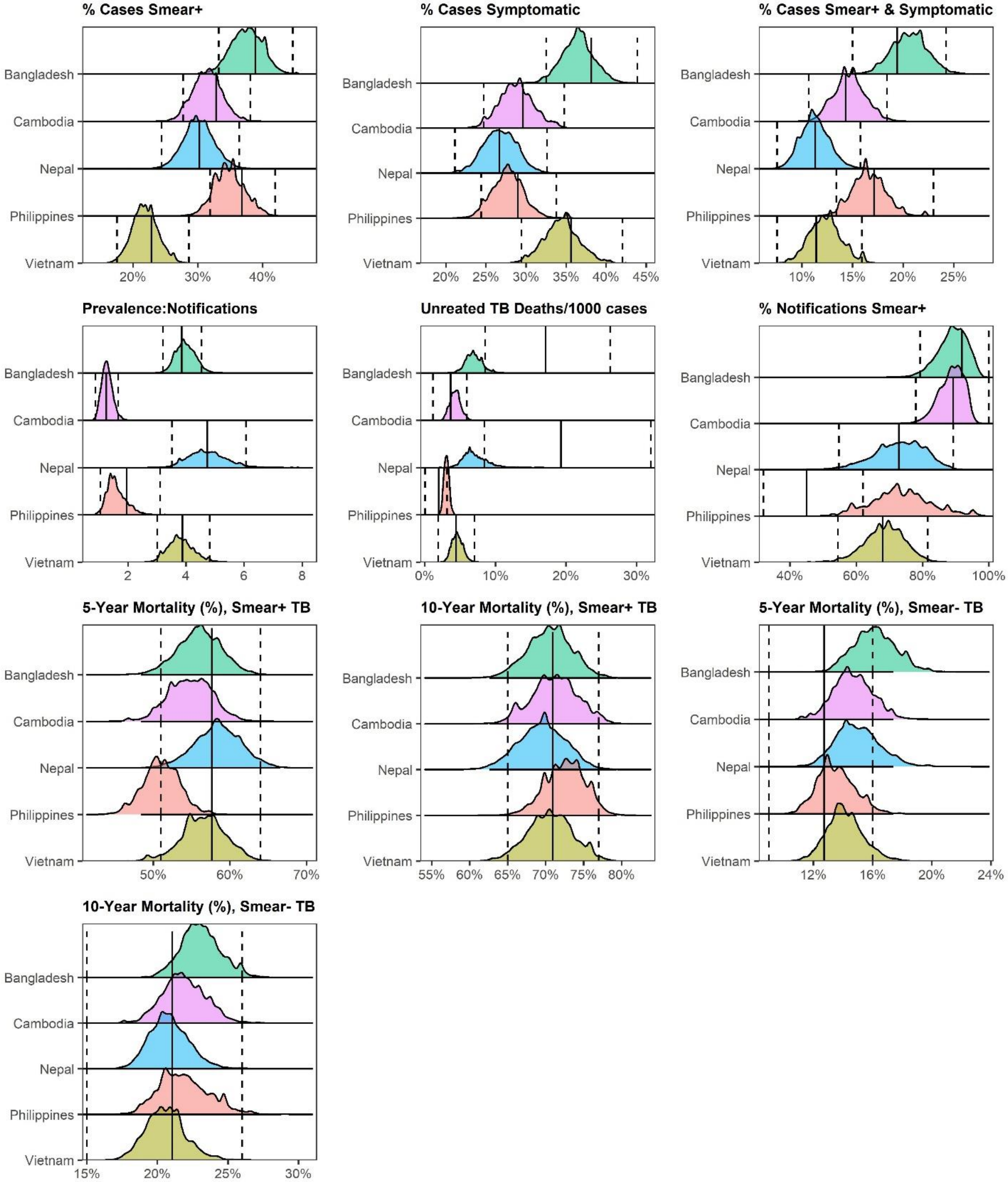

Curves show densities of modeled outputs generated using posterior parameters for each country. Calibration target means and 95% confidence intervals are indicated by solid and dashed vertical lines, respectively.

**Figure E3: Calibration performance: distribution of likelihoods across 50 IMIS chains per country**

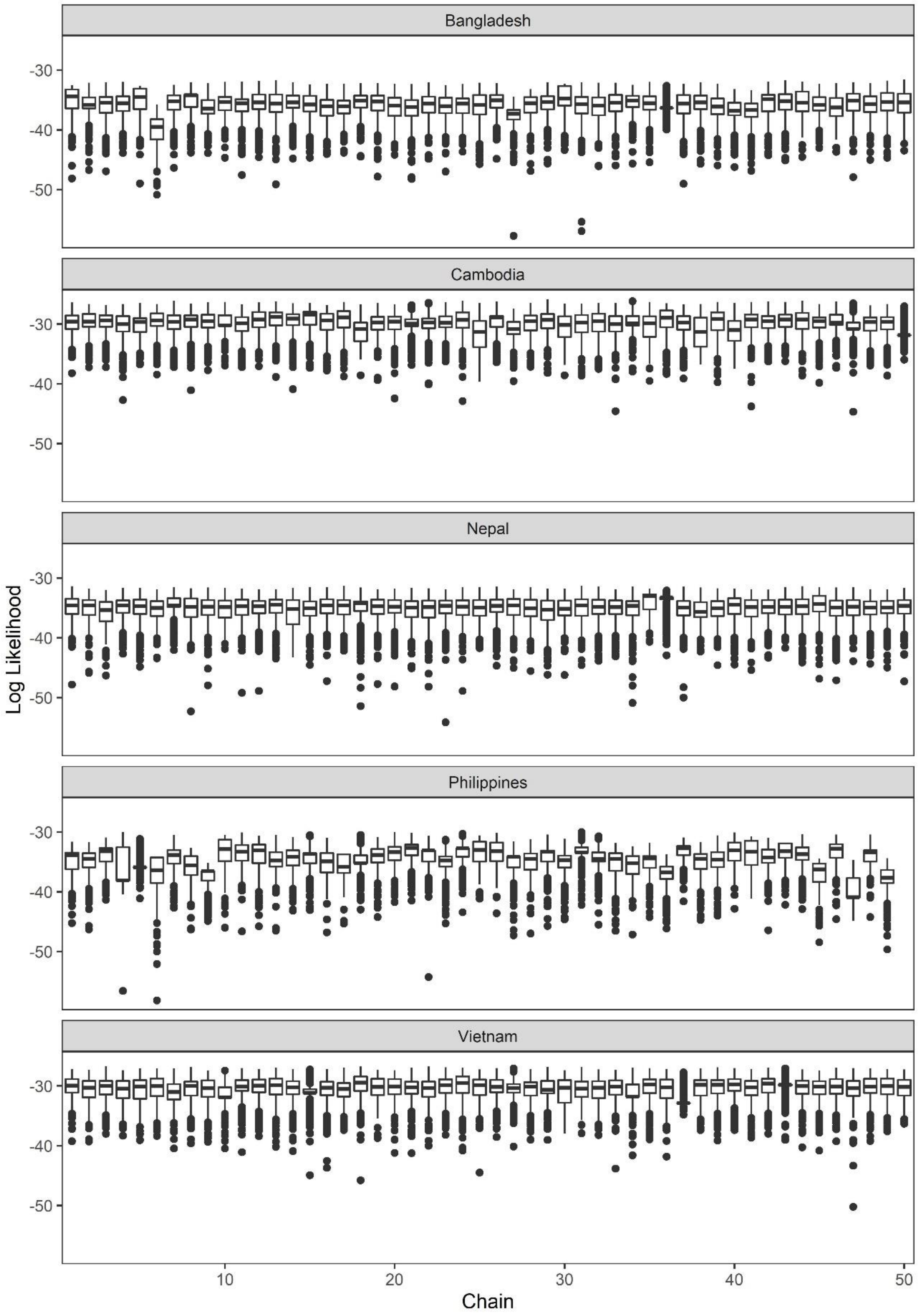

Figure shows the means (horizontal lines within boxes), 25% and 75% quantiles (boxes), 2.5th and 97.5th percentiles (whiskers) and outliers (dots) of the log likelihood from the 1000 posterior parameters sampled from each of the 50 IMIS chains for each country.

**Figure E4: Individuals TB natural history trajectory characteristics over five years by country – durations with TB**

**Bangladesh**

| | Smear-Negative Subclinical | Smear-Positive Subclinical | Smear-Negative Symptomatic | Smear-Positive Symptomatic | Total Population |
|---|---|---|---|---|---|
| Smear-Negative Subclinical | 2.1 [1.8-2.7] | 0.3 [0.1-0.8] | 1.8 [1.5-2.3] | 0.2 [0.0-0.3] | 1.3 [1.1-1.7] |
| Smear-Positive Subclinical | 0.7 [0.5-1.1] | 7.7 [5.2-10.4] | 1.0 [0.5-1.6] | 3.9 [1.0-6.6] | 2.8 [1.6-3.9] |
| Smear-Negative Symptomatic | 0.7 [0.5-1.0] | 0.2 [0.0-0.3] | 2.6 [2.2-3.3] | 0.1 [0.0-0.2] | 0.8 [0.7-1.1] |
| Smear-Positive Symptomatic | 0.9 [0.7-1.2] | 7.8 [5.7-10.0] | 1.6 [1.3-2.1] | 8.4 [6.6-10.6] | 3.8 [3.0-4.8] |
| Total with TB (any) | 4.4 [3.7-5.7] | 16.0 [12.4-19.4] | 7.1 [6.1-8.5] | 12.6 [9.5-15.8] | 8.7 [7.2-10.4] |

**Cambodia**

| | Smear-Negative Subclinical | Smear-Positive Subclinical | Smear-Negative Symptomatic | Smear-Positive Symptomatic | Total Population |
|---|---|---|---|---|---|
| Smear-Negative Subclinical | 2.2 [1.8-3.4] | 0.3 [0.1-0.9] | 1.9 [1.5-3.0] | 0.1 [0.0-0.2] | 1.5 [1.2-2.4] |
| Smear-Positive Subclinical | 0.7 [0.4-1.0] | 7.9 [5.5-10.4] | 0.8 [0.5-1.3] | 2.6 [0.5-5.4] | 2.3 [1.5-3.2] |
| Smear-Negative Symptomatic | 0.6 [0.4-1.0] | 0.1 [0.0-0.2] | 2.6 [2.1-3.4] | 0.1 [0.0-0.1] | 0.7 [0.5-1.0] |
| Smear-Positive Symptomatic | 0.6 [0.4-1.0] | 5.7 [4.0-8.0] | 1.1 [0.8-1.6] | 6.3 [4.5-8.7] | 2.4 [1.8-3.3] |
| Total with TB (any) | 4.1 [3.3-6.3] | 14.0 [11.1-17.6] | 6.5 [5.4-8.6] | 9.1 [6.4-12.6] | 7.0 [5.7-9.1] |

**Nepal**

| | Smear-Negative Subclinical | Smear-Positive Subclinical | Smear-Negative Symptomatic | Smear-Positive Symptomatic | Total Population |
|---|---|---|---|---|---|
| Smear-Negative Subclinical | 2.3 [1.8-3.6] | 0.4 [0.1-0.8] | 2.0 [1.5-3.1] | 0.3 [0.1-0.5] | 1.7 [1.3-2.6] |
| Smear-Positive Subclinical | 0.8 [0.5-1.3] | 10.4 [7.2-14.1] | 1.2 [0.7-1.7] | 6.3 [2.9-9.8] | 3.3 [2.2-4.5] |
| Smear-Negative Symptomatic | 0.7 [0.4-1.1] | 0.2 [0.1-0.4] | 2.6 [2.1-3.4] | 0.2 [0.1-0.3] | 0.8 [0.6-1.1] |
| Smear-Positive Symptomatic | 0.5 [0.3-0.7] | 5.4 [3.6-7.7] | 0.9 [0.6-1.2] | 5.8 [4.0-8.2] | 2.1 [1.4-2.9] |
| Total with TB (any) | 4.2 [3.3-6.5] | 16.5 [12.5-21.2] | 6.6 [5.5-8.7] | 12.6 [8.7-17.1] | 7.8 [6.2-10.2] |

**Philippines**

| | Smear-Negative Subclinical | Smear-Positive Subclinical | Smear-Negative Symptomatic | Smear-Positive Symptomatic | Total Population |
|---|---|---|---|---|---|
| Smear-Negative Subclinical | 2.5 [1.8-4.0] | 0.4 [0.0-1.0] | 1.9 [1.2-3.2] | 0.1 [0.0-0.1] | 1.6 [1.1-2.5] |
| Smear-Positive Subclinical | 0.8 [0.5-1.3] | 9.8 [6.9-14.2] | 0.8 [0.5-1.2] | 1.5 [0.1-4.9] | 2.7 [1.8-4.0] |
| Smear-Negative Symptomatic | 0.5 [0.3-0.9] | 0.1 [0.0-0.2] | 2.8 [2.0-4.0] | 0.0 [0.0-0.1] | 0.6 [0.4-0.9] |
| Smear-Positive Symptomatic | 0.8 [0.5-1.3] | 7.3 [5.0-10.2] | 1.4 [0.9-2.0] | 8.5 [5.7-12.2] | 3.4 [2.5-4.7] |
| Total with TB (any) | 4.6 [3.3-7.2] | 17.6 [13.0-23.5] | 6.8 [5.2-9.2] | 10.0 [6.8-14.3] | 8.3 [6.4-11.2] |

**Vietnam**

| | Smear-Negative Subclinical | Smear-Positive Subclinical | Smear-Negative Symptomatic | Smear-Positive Symptomatic | Total Population |
|---|---|---|---|---|---|
| Smear-Negative Subclinical | 2.4 [1.9-3.9] | 0.3 [0.1-0.8] | 2.0 [1.5-3.3] | 0.2 [0.0-0.3] | 1.8 [1.4-2.9] |
| Smear-Positive Subclinical | 0.4 [0.2-0.8] | 6.3 [3.8-9.7] | 0.6 [0.3-1.0] | 2.8 [0.7-5.4] | 1.4 [0.7-2.2] |
| Smear-Negative Symptomatic | 1.0 [0.6-1.5] | 0.2 [0.1-0.4] | 3.2 [2.4-4.5] | 0.2 [0.0-0.3] | 1.3 [1.0-1.9] |
| Smear-Positive Symptomatic | 0.5 [0.3-0.9] | 6.3 [4.5-8.6] | 1.0 [0.7-1.4] | 6.7 [4.9-9.1] | 2.0 [1.5-2.7] |
| Total with TB (any) | 4.3 [3.2-6.7] | 13.1 [10.1-16.9] | 6.8 [5.5-9.1] | 9.9 [7.0-13.3] | 6.5 [5.3-8.6] |

4 8 12 16

Figure shows the average cumulative durations (in months) spent in each specified state (rows), conditional on being in each starting state (columns) at time 0, by country.

**Figure E5: Individuals TB natural history trajectory characteristics over five years by country – proportions reaching TB-related outcomes**

**Bangladesh**

| | Smear-Negative Subclinical | Smear-Positive Subclinical | Smear-Negative Symptomatic | Smear-Positive Symptomatic | Total Population |
|---|---|---|---|---|---|
| Any smear-positive TB state | 11% [9-16%] | NA | 20% [16-26%] | NA | 47% [46-50%] |
| Any symptomatic TB state | 32% [25-39%] | 94% [80-99%] | NA | NA | 70% [65-74%] |
| Spontaneous resolution | 89% [85-91%] | 14% [3-32%] | 77% [72-81%] | 8% [2-13%] | 56% [52-61%] |
| Treatment | 8% [7-11%] | 65% [50-78%] | 17% [14-22%] | 70% [60-81%] | 33% [28-38%] |
| TB Mortality | 3% [2-4%] | 19% [13-24%] | 5% [3-8%] | 21% [15-26%] | 10% [7-12%] |

**Cambodia**

| | Smear-Negative Subclinical | Smear-Positive Subclinical | Smear-Negative Symptomatic | Smear-Positive Symptomatic | Total Population |
|---|---|---|---|---|---|
| Any smear-positive TB state | 10% [7-15%] | NA | 19% [14-24%] | NA | 41% [39-44%] |
| Any symptomatic TB state | 28% [19-41%] | 92% [75-99%] | NA | NA | 61% [55-69%] |
| Spontaneous resolution | 89% [84-93%] | 13% [3-33%] | 77% [71-82%] | 4% [1-9%] | 61% [57-66%] |
| Treatment | 9% [6-13%] | 73% [56-85%] | 19% [14-25%] | 81% [72-89%] | 33% [28-37%] |
| TB Mortality | 1% [1-2%] | 12% [7-17%] | 3% [2-5%] | 14% [9-19%] | 5% [4-8%] |

**Nepal**

| | Smear-Negative Subclinical | Smear-Positive Subclinical | Smear-Negative Symptomatic | Smear-Positive Symptomatic | Total Population |
|---|---|---|---|---|---|
| Any smear-positive TB state | 9% [6-13%] | NA | 15% [11-20%] | NA | 37% [35-40%] |
| Any symptomatic TB state | 30% [21-42%] | 95% [85-99%] | NA | NA | 61% [55-67%] |
| Spontaneous resolution | 90% [85-93%] | 16% [5-31%] | 77% [71-82%] | 11% [4-20%] | 65% [61-70%] |
| Treatment | 7% [5-11%] | 58% [40-74%] | 17% [12-23%] | 62% [45-77%] | 24% [18-31%] |
| TB Mortality | 2% [1-4%] | 23% [15-32%] | 5% [3-8%] | 25% [16-35%] | 9% [6-13%] |

**Philippines**

| | Smear-Negative Subclinical | Smear-Positive Subclinical | Smear-Negative Symptomatic | Smear-Positive Symptomatic | Total Population |
|---|---|---|---|---|---|
| Any smear-positive TB state | 10% [7-16%] | NA | 17% [13-23%] | NA | 44% [42-48%] |
| Any symptomatic TB state | 24% [15-39%] | 90% [72-99%] | NA | NA | 65% [58-73%] |
| Spontaneous resolution | 88% [79-92%] | 13% [0-33%] | 68% [55-80%] | 2% [0-5%] | 56% [49-62%] |
| Treatment | 11% [6-18%] | 74% [54-88%] | 29% [17-43%] | 85% [77-91%] | 38% [31-45%] |
| TB Mortality | 1% [1-2%] | 10% [7-15%] | 2% [2-4%] | 12% [8-17%] | 5% [3-7%] |

**Vietnam**

| | Smear-Negative Subclinical | Smear-Positive Subclinical | Smear-Negative Symptomatic | Smear-Positive Symptomatic | Total Population |
|---|---|---|---|---|---|
| Any smear-positive TB state | 8% [6-12%] | NA | 15% [11-20%] | NA | 31% [29-34%] |
| Any symptomatic TB state | 33% [24-46%] | 95% [84-99%] | NA | NA | 64% [59-71%] |
| Spontaneous resolution | 89% [84-93%] | 12% [3-27%] | 75% [69-81%] | 6% [2-11%] | 68% [63-71%] |
| Treatment | 9% [6-13%] | 71% [58-84%] | 21% [16-27%] | 76% [66-86%] | 26% [22-31%] |
| TB Mortality | 2% [1-3%] | 16% [10-22%] | 4% [2-5%] | 17% [11-23%] | 5% [4-7%] |

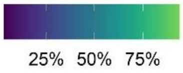

Figure shows the proportion of those in each starting state at time 0 (columns) ever reaching other states/outcomes (rows), by country.

### Table E2: Per-person and population contribution of TB states to relative secondary infections

| | Bangladesh | Cambodia | Nepal | Philippines | Vietnam | Pooled |
|---|---|---|---|---|---|---|
| **Per-person contribution to transmission over 5 years, vs. smear-negative subclinical at time 0** | | | | | | |
| Smear-positive subclinical | 6.2 [4.5- 7.8] | 6.2 [4.2- 8.5] | 7.3 [4.7-10.0] | 7.0 [4.2-10.1] | 6.4 [4.1- 8.9] | 6.7 [4.4- 9.4] |
| Smear-negative symptomatic | 1.7 [1.5- 2.0] | 1.7 [1.4- 2.1] | 1.7 [1.4- 2.1] | 1.6 [1.2- 2.1] | 1.8 [1.4- 2.2] | 1.7 [1.4- 2.1] |
| Smear-positive symptomatic | 5.2 [3.8- 6.7] | 4.5 [3.0- 6.5] | 5.9 [3.7- 8.3] | 4.7 [2.7- 6.9] | 5.2 [3.3- 7.6] | 5.2 [3.2- 7.6] |
| **Per-person contribution to transmission over 5 years, vs. smear-positive symptomatic at time 0** | | | | | | |
| Smear-positive subclinical | 1.2 [1.1- 1.3] | 1.4 [1.2- 1.7] | 1.3 [1.1- 1.4] | 1.5 [1.2- 1.8] | 1.2 [1.1- 1.4] | 1.3 [1.1- 1.7] |
| **Proportion of prevalent TB by state (based on the prevalence survey data)** | | | | | | |
| Smear-negative subclinical | 42% [37-48%] | 52% [46-57%] | 54% [48-61%] | 51% [46-56%] | 53% [47-59%] | NA |
| Smear-positive subclinical | 19% [15-24%] | 18% [14-23%] | 19% [14-24%] | 20% [16-24%] | 11% [7-16%] | NA |
| Smear-negative symptomatic | 19% [14-23%] | 15% [11-19%] | 15% [11-20%] | 12% [9-15%] | 24% [19-30%] | NA |
| Smear-positive symptomatic | 19% [15-24%] | 14% [11-18%] | 11% [8-16%] | 17% [13-21%] | 11% [7-16%] | NA |
| **Population contribution cumulative 5-year transmission, by state at time 0** | | | | | | |
| Smear-negative subclinical | 9% [7-11%] | 12% [9-16%] | 11% [8-15%] | 10% [7-15%] | 14% [10-19%] | NA |
| Smear-positive subclinical | 42% [39-45%] | 44% [38-49%] | 43% [39-48%] | 48% [42-53%] | 37% [31-41%] | NA |
| Smear-negative symptomatic | 13% [11-16%] | 15% [13-19%] | 14% [11-18%] | 12% [10-16%] | 20% [16-25%] | NA |
| Smear-positive symptomatic | 36% [32-39%] | 29% [24-34%] | 32% [27-37%] | 30% [25-35%] | 29% [23-34%] | NA |

The first 2 sections of the table show the relative number of secondary infections generated over a five-year period by someone starting out in various TB states at time 0, compared to someone starting out smear-negative and subclinical (section A) or smear-positive and symptomatic (section B) at time 0. Section C of the table shows the breakdown of prevalence by smear and symptom status (based on data from the Philippines prevalence survey) Section D shows the percent of cumulative transmission over 5 years generated by a population with prevalent TB at time 0 attributable to each of the 4 initial TB states, accounting for the population-level prevalence of each TB state. Pooled estimates were generated by taking means, 2.5$^{th}$ percentiles, and 97.5$^{th}$ percentiles across 100,000 simulations for each country.

**Figure E6: Posterior parameter distributions from sensitivity analyses**

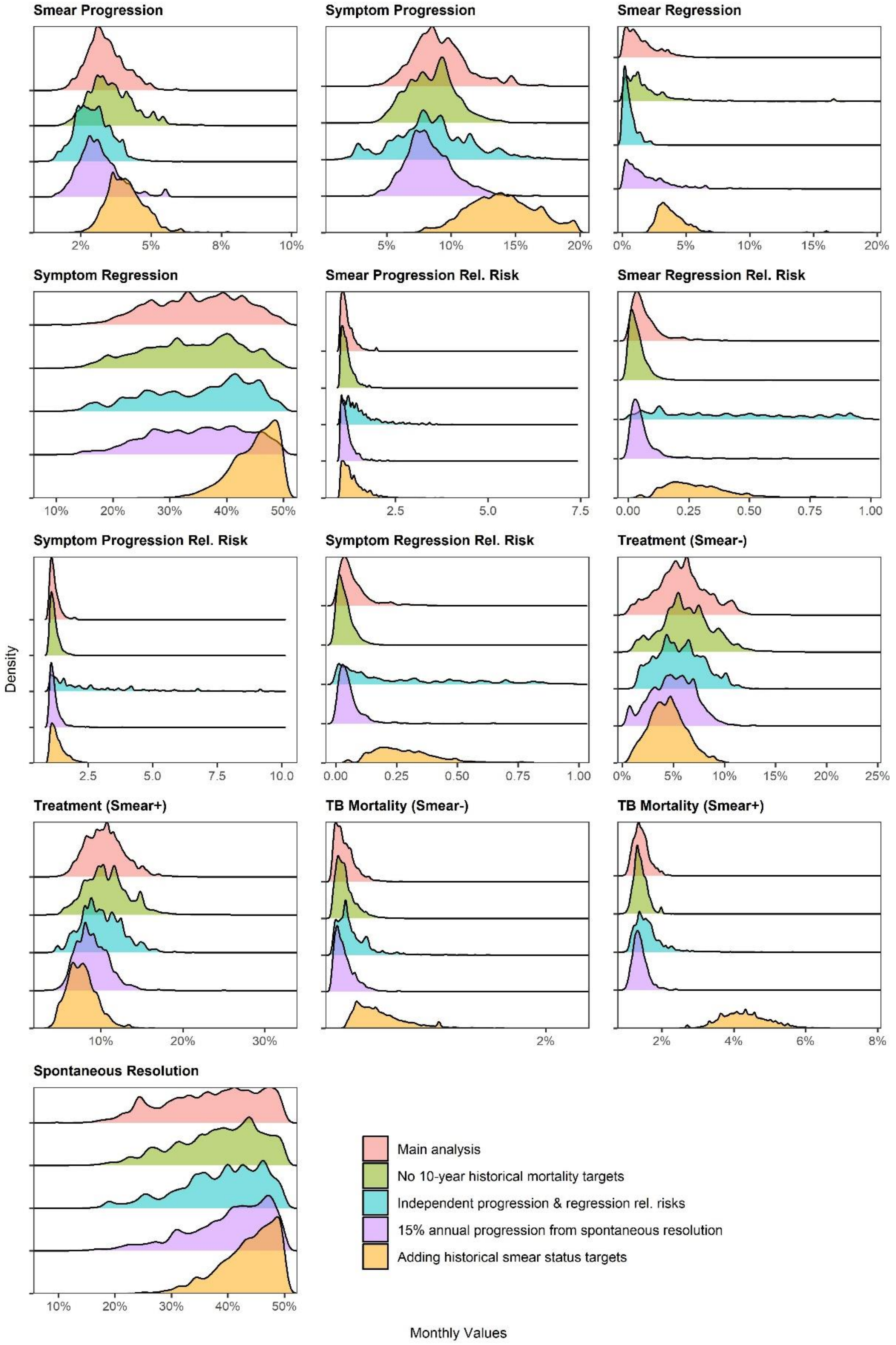

Curves show posterior parameter distributions for each sensitivity analysis. Except for the third sensitivity analysis (independent progression & regression relative risks; shown in aqua), symptom progression relative risks were set to equal smear progression relative risks and symptom regression relative risks were set to equal smear regression relative risks. All sensitivity analyses were conducted using data from the Philippines.

**Figure E7: Fit to targets from sensitivity analyses**

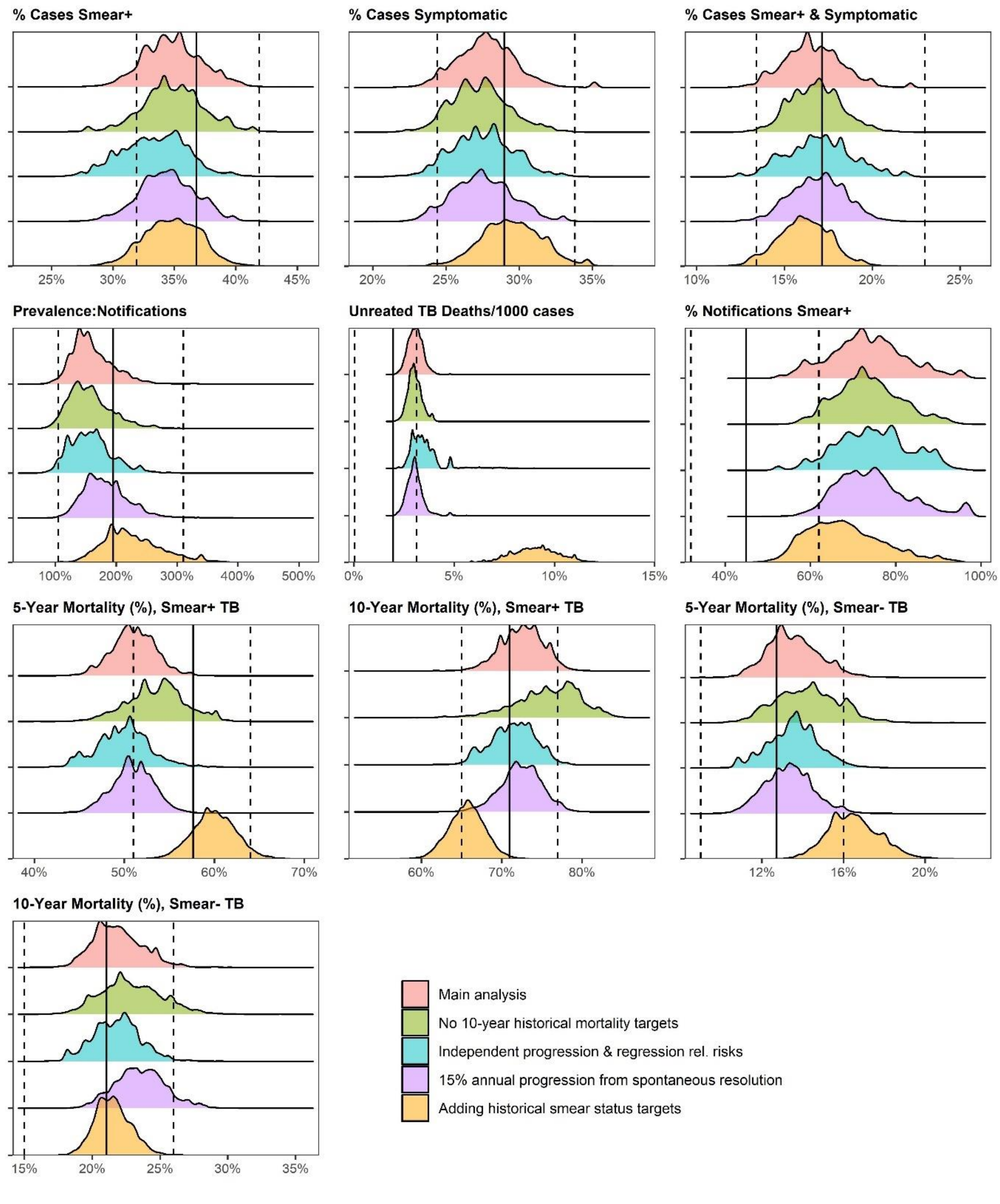

Curves show densities of modeled outputs generated using posterior parameters for each sensitivity analysis. Calibration target means and 95% confidence intervals for the main calibration targets are indicated by solid and dashed vertical lines, respectively. All sensitivity analyses were conducted using data from the Philippines.

**Figure E8: Individuals TB natural history trajectory characteristics over five years from sensitivity analyses – durations with TB**

**Main analysis**

| | Smear-Negative Subclinical | Smear-Positive Subclinical | Smear-Negative Symptomatic | Smear-Positive Symptomatic | Total Population |
|---|---|---|---|---|---|
| Smear-Negative Subclinical | 2.5 [1.8-4.0] | 0.4 [0.0-1.0] | 1.9 [1.2-3.2] | 0.1 [0.0-0.1] | 1.6 [1.1-2.5] |
| Smear-Positive Subclinical | 0.8 [0.5-1.3] | 9.8 [6.9-14.2] | 0.8 [0.5-1.2] | 1.5 [0.1-4.9] | 2.7 [1.8-4.0] |
| Smear-Negative Symptomatic | 0.5 [0.3-0.9] | 0.1 [0.0-0.2] | 2.8 [2.0-4.0] | 0.0 [0.0-0.1] | 0.6 [0.4-0.9] |
| Smear-Positive Symptomatic | 0.8 [0.5-1.3] | 7.3 [5.0-10.2] | 1.4 [0.9-2.0] | 8.5 [5.7-12.2] | 3.4 [2.5-4.7] |
| **Total with TB (any)** | 4.6 [3.3-7.2] | 17.6 [13.0-23.5] | 6.8 [5.2-9.2] | 10.0 [6.8-14.3] | 8.3 [6.4-11.2] |

**Independent progression & regression rel. risks**

| | Smear-Negative Subclinical | Smear-Positive Subclinical | Smear-Negative Symptomatic | Smear-Positive Symptomatic | Total Population |
|---|---|---|---|---|---|
| Smear-Negative Subclinical | 2.4 [1.8-4.1] | 0.2 [0.0-0.5] | 1.9 [1.2-3.2] | 0.1 [0.0-0.2] | 1.5 [1.1-2.5] |
| Smear-Positive Subclinical | 0.7 [0.4-1.2] | 9.2 [5.1-13.4] | 0.9 [0.3-1.4] | 3.4 [0.1-7.9] | 2.9 [1.5-4.2] |
| Smear-Negative Symptomatic | 0.5 [0.2-0.8] | 0.1 [0.0-0.2] | 2.6 [1.9-4.4] | 0.0 [0.0-0.2] | 0.6 [0.4-0.8] |
| Smear-Positive Symptomatic | 0.7 [0.5-1.2] | 8.1 [5.5-12.2] | 1.4 [0.9-2.1] | 8.5 [5.7-13.1] | 3.6 [2.5-5.2] |
| **Total with TB (any)** | 4.3 [3.1-6.9] | 17.5 [12.5-23.7] | 6.8 [5.2-9.5] | 11.9 [7.5-18.7] | 8.5 [6.2-11.5] |

**15% annual progression from spontaneous resolution**

| | Smear-Negative Subclinical | Smear-Positive Subclinical | Smear-Negative Symptomatic | Smear-Positive Symptomatic | Total Population |
|---|---|---|---|---|---|
| Smear-Negative Subclinical | 3.8 [3.0-6.0] | 0.6 [0.0-1.8] | 2.9 [2.1-4.9] | 0.1 [0.0-0.2] | 2.4 [1.9-3.8] |
| Smear-Positive Subclinical | 1.1 [0.7-1.8] | 10.3 [6.3-15.1] | 1.0 [0.6-1.7] | 1.6 [0.1-6.1] | 3.0 [1.8-4.5] |
| Smear-Negative Symptomatic | 0.7 [0.4-1.2] | 0.1 [0.0-0.3] | 3.0 [2.1-4.7] | 0.0 [0.0-0.1] | 0.7 [0.5-1.1] |
| Smear-Positive Symptomatic | 1.0 [0.7-1.6] | 8.0 [4.8-11.3] | 1.6 [1.1-2.3] | 9.6 [6.6-13.3] | 3.9 [2.8-5.3] |
| **Total with TB (any)** | 6.5 [5.3-10.3] | 19.0 [13.5-25.8] | 8.5 [6.7-11.9] | 11.3 [7.8-16.4] | 10.0 [7.9-13.4] |

**Adding historical smear status targets**

| | Smear-Negative Subclinical | Smear-Positive Subclinical | Smear-Negative Symptomatic | Smear-Positive Symptomatic | Total Population |
|---|---|---|---|---|---|
| Smear-Negative Subclinical | 3.4 [2.9-4.3] | 1.1 [0.7-1.8] | 2.8 [2.2-3.6] | 0.6 [0.4-0.8] | 2.4 [2.0-3.0] |
| Smear-Positive Subclinical | 1.1 [0.8-1.5] | 8.0 [5.7-10.0] | 1.3 [0.8-1.8] | 3.9 [1.7-6.3] | 3.0 [2.0-3.9] |
| Smear-Negative Symptomatic | 0.9 [0.6-1.3] | 0.4 [0.3-0.5] | 2.5 [2.1-3.1] | 0.3 [0.2-0.4] | 0.9 [0.6-1.2] |
| Smear-Positive Symptomatic | 0.9 [0.7-1.2] | 5.8 [4.2-7.4] | 1.4 [1.1-1.9] | 6.9 [5.2-9.2] | 3.0 [2.3-3.7] |
| **Total with TB (any)** | 6.3 [5.4-8.0] | 15.2 [11.8-18.1] | 8.0 [6.8-9.6] | 11.7 [8.8-15.1] | 9.2 [7.7-10.8] |

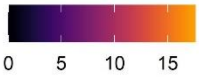

Figure shows the average cumulative durations (in months) spent in each specified state (rows), conditional on being in each starting state (columns) at time 0, by sensitivity analysis. All sensitivity analyses were run using data from the Philippines.

**Figure E9: Individuals TB natural history trajectory characteristics over five years from sensitivity analyses – proportions reaching TB-related outcomes**

**Main analysis**

| | Smear-Negative Subclinical | Smear-Positive Subclinical | Smear-Negative Symptomatic | Smear-Positive Symptomatic | Total Population |
|---|---|---|---|---|---|
| Any smear-positive TB state | 10% [7-16%] | NA | 17% [13-23%] | NA | 44% [42-48%] |
| Any symptomatic TB state | 24% [15-39%] | 90% [72-99%] | NA | NA | 65% [58-73%] |
| Spontaneous resolution | 88% [79-92%] | 13% [0-33%] | 68% [55-80%] | 2% [0-5%] | 56% [49-62%] |
| Treatment | 11% [6-18%] | 74% [54-88%] | 29% [17-43%] | 85% [77-91%] | 38% [31-45%] |
| TB Mortality | 1% [1-2%] | 10% [7-15%] | 2% [2-4%] | 12% [8-17%] | 5% [3-7%] |

**Independent progression & regression rel. risks**

| | Smear-Negative Subclinical | Smear-Positive Subclinical | Smear-Negative Symptomatic | Smear-Positive Symptomatic | Total Population |
|---|---|---|---|---|---|
| Any smear-positive TB state | 9% [6-14%] | NA | 17% [12-22%] | NA | 43% [41-46%] |
| Any symptomatic TB state | 20% [12-35%] | 91% [73-99%] | NA | NA | 67% [59-75%] |
| Spontaneous resolution | 89% [83-93%] | 6% [0-16%] | 69% [57-81%] | 2% [0-7%] | 55% [51-60%] |
| Treatment | 10% [5-15%] | 78% [63-88%] | 27% [15-40%] | 82% [66-90%] | 37% [30-44%] |
| TB Mortality | 1% [1-2%] | 14% [8-23%] | 3% [2-5%] | 14% [8-23%] | 6% [4-10%] |

**15% annual progression from spontaneous resolution**

| | Smear-Negative Subclinical | Smear-Positive Subclinical | Smear-Negative Symptomatic | Smear-Positive Symptomatic | Total Population |
|---|---|---|---|---|---|
| Any smear-positive TB state | 14% [9-21%] | NA | 19% [14-26%] | NA | 46% [43-51%] |
| Any symptomatic TB state | 32% [22-48%] | 88% [64-99%] | NA | NA | 63% [55-72%] |
| Spontaneous resolution | 90% [84-94%] | 16% [1-47%] | 72% [61-82%] | 2% [0-5%] | 58% [53-66%] |
| Treatment | 12% [7-19%] | 70% [46-86%] | 28% [17-40%] | 83% [73-90%] | 38% [30-45%] |
| TB Mortality | 1% [1-2%] | 11% [7-17%] | 3% [2-4%] | 13% [9-19%] | 6% [4-8%] |

**Adding historical smear status targets**

| | Smear-Negative Subclinical | Smear-Positive Subclinical | Smear-Negative Symptomatic | Smear-Positive Symptomatic | Total Population |
|---|---|---|---|---|---|
| Any smear-positive TB state | 17% [13-23%] | NA | 24% [19-30%] | NA | 48% [46-52%] |
| Any symptomatic TB state | 42% [31-54%] | 89% [76-96%] | NA | NA | 68% [60-75%] |
| Spontaneous resolution | 89% [85-93%] | 30% [22-43%] | 74% [66-81%] | 17% [12-22%] | 64% [59-69%] |
| Treatment | 11% [6-17%] | 45% [32-55%] | 22% [13-32%] | 53% [40-66%] | 26% [19-34%] |
| TB Mortality | 4% [3-6%] | 25% [15-34%] | 7% [5-10%] | 30% [22-38%] | 13% [9-17%] |

25% 50% 75%

Figure shows the proportion of those in each starting state at time 0 (columns) ever reaching other states/outcomes (rows), by country. All sensitivity analyses were run using data from the Philippines.

**Table E3: Per-person and population contribution of TB states to relative secondary infections - sensitivity analyses**

| | Main Model (Philippines) | Analysis decoupling progression & regression relative risks | Analysis with non-zero progression from spontaneous resolution | Analysis that includes historical smear regression targets |
|---|---|---|---|---|
| **A. Per-person contribution to transmission over 5 years, vs. smear-negative subclinical at time 0** | | | | |
| Smear-positive subclinical | 7.0 [4.2-10.0] | 7.8 [4.9-11.1] | 5.4 [3.4-7.9] | 4.1 [3.2-5.1] |
| Smear-negative symptomatic | 1.6 [1.2-2.1] | 1.8 [1.3-2.4] | 1.4 [1.1-2.8] | 1.4 [1.2-1.6] |
| Smear-positive symptomatic | 4.7 [2.7-6.9] | 5.8 [3.3-9.2] | 3.7 [2.4-5.6] | 3.5 [2.6-4.5] |
| **B. Per-person contribution to transmission over 5 years, vs. smear-positive symptomatic at time 0** | | | | |
| Smear-positive subclinical | 1.5 [1.2-1.8] | 1.4 [1.1-1.8] | 1.4 [1.1-1.8] | 1.2 [1.1-1.3] |
| **C. Proportion of prevalent TB by state (based on the prevalence survey data)** | | | | |
| Smear-negative subclinical | 51% [46-56%] | 51% [46-56%] | 51% [46-56%] | 51% [46-56%] |
| Smear-positive subclinical | 20% [16-24%] | 20% [16-24%] | 20% [16-24%] | 20% [16-24%] |
| Smear-negative symptomatic | 12% [9-15%] | 12% [9-15%] | 12% [9-15%] | 12% [9-15%] |
| Smear-positive symptomatic | 17% [13-21%] | 17% [13-21%] | 17% [13-21%] | 17% [13-21%] |
| **D. Population contribution cumulative 5-year transmission, by state at time 0** | | | | |
| Smear-negative subclinical | 10% [7-15%] | 9% [7-13%] | 13% [9-17%] | 14% [11-17%] |
| Smear-positive subclinical | 48% [42-53%] | 46% [41-53%] | 44% [38-50%] | 39% [36-42%] |
| Smear-negative symptomatic | 12% [9-16%] | 12% [9-15%] | 14% [11-18%] | 15% [13-18%] |
| Smear-positive symptomatic | 30% [25-35%] | 33% [26-39%] | 29% [24-35%] | 32% [28-35%] |

All sensitivity analyses were run using data from the Philippines. The first 2 sections of the table show the relative number of secondary infections generated over a five-year period by someone starting out in various TB states at time 0, compared to someone starting out smear-negative and subclinical (section A) or smear-positive and symptomatic (section B) at time 0. Section C of the table shows the breakdown of prevalence by smear and symptom status (based on data from the Philippines prevalence survey) Section D shows the percent of cumulative transmission over 5 years generated by a population with prevalent TB at time 0 attributable to each of the 4 initial TB states, accounting for the population-level prevalence of each TB state.
